# Risk factors for developing COVID-19: a population-based longitudinal study (COVIDENCE UK)

**DOI:** 10.1101/2021.03.27.21254452

**Authors:** Hayley Holt, Mohammad Talaei, Matthew Greenig, Dominik Zenner, Jane Symons, Clare Relton, Katherine S Young, Molly R Davies, Katherine N Thompson, Jed Ashman, Sultan Saeed Rajpoot, Ahmed Ali Kayyale, Sarah El Rifai, Philippa J. Lloyd, David A. Jolliffe, Sarah Finer, Stamatina Ilidriomiti, Alec Miners, Nicholas S. Hopkinson, Bodrul Alam, Paul E Pfeffer, David McCoy, Gwyneth A Davies, Ronan A Lyons, Christopher J Griffiths, Frank Kee, Aziz Sheikh, Gerome Breen, Seif O Shaheen, Adrian R Martineau

**Affiliations:** Institute of Population Health Sciences, Barts and The London School of Medicine and Dentistry, Queen Mary University of London, London, UK; Asthma UK Centre for Applied Research, Queen Mary University of London, London, UK; Blizard Institute, Barts and The London School of Medicine and Dentistry, Queen Mary University of London, London, UK; Jane Symons Media, London; Institute of Psychiatry, Psychology and Neuroscience, King’s College London, London, UK; NIHR Maudsley Biomedical Research Centre, King’s College London, UK; Department of Health Services Research and Policy, London School of Hygiene and Tropical Medicine, London, UK; National Heart and Lung Institute, Imperial College London, UK; Edenfield Road Surgery, Rochdale, UK; Population Data Science, Swansea University Medical School, Singleton Park, Swansea, UK; Centre for Public Health Research (NI), Queen’s University Belfast, Belfast, UK; Usher Institute, University of Edinburgh, Edinburgh, UK

## Abstract

**Background:** Risk factors for severe COVID-19 include older age, male sex, obesity, Black or Asian ethnicity and underlying medical conditions. Whether these factors also influence susceptibility to developing COVID-19 is uncertain.

**Methods:** We undertook a prospective, population-based cohort study (COVIDENCE UK) from 1^st^ May 2020 to 5^th^ February 2021. Baseline information on potential risk factors was captured by an online questionnaire. Monthly follow-up questionnaires captured incident COVID-19. We used logistic regression models to estimate multivariable-adjusted odds ratios (aORs) for associations between potential risk factors and risk of COVID-19.

**Findings:** We recorded 446 incident cases of COVID-19 in 15,227 participants (2.9%). Increased risk of developing COVID-19 was independently associated with Asian/Asian British *vs*. White ethnicity (aOR 2.31, 95% CI 1.35-3.95), household overcrowding (aOR per additional 0.5 people/bedroom 1.26, 1.11-1.43), any *vs*. no visits to/from other households in previous week (aOR 1.33, 1.07-1.64), number of visits to indoor public places (aOR per extra visit per week 1.05, 1.01-1.09), frontline occupation excluding health/social care *vs*. no frontline occupation (aOR 1.49, 1.12-1.98), and raised body mass index (BMI) (aOR 1.51 [1.20-1.90] for BMI 25.0-30.0 kg/m^2^ and 1.38 [1.05-1.82] for BMI >30.0 kg/m^2^ *vs*. BMI <25.0 kg/m^2^). Atopic disease was independently associated with decreased risk (aOR 0.76, 0.59-0.98). No independent associations were seen for age, sex, other medical conditions, diet, or micronutrient supplement use.

**Interpretation:** After rigorous adjustment for factors influencing exposure to SARS-CoV-2, Asian/Asian British ethnicity and raised BMI were associated with increased risk of developing COVID-19, while atopic disease was associated with decreased risk.

**Funding:** Barts Charity, Health Data Research UK

## Introduction

COVID-19 has taken a heavy toll on the health of populations globally.^1-3^ Risk factors for severe and fatal disease include male sex, Black or Asian ethnic origin, obesity, deprivation and a range of comorbidities including diabetes mellitus, cardiovascular disease, chronic obstructive pulmonary disease (COPD) and hypertension.^4,5^ However, there is growing evidence from population-based studies to suggest that at least some risk factors for developing COVID-19 -irrespective of severity - may be distinct from those which predispose to disease at the most severe end of the spectrum. For example, population-based studies in both the US and the UK have reported that risk of COVID-19 is higher in younger vs. older adults ^6^, a finding supported by serology studies in the UK and Switzerland reporting higher prevalence of antibodies to SARS-CoV-2 in younger vs older adults.^7,8^ Again, in contrast to studies reporting that diabetes, heart disease and hypertension are risk factors for severe disease, the presence of pre-existing health conditions has been reported to associate with decreased, rather than increased, risk of SARS-CoV-2 sero-positivity in a population-based study conducted in the UK.^7^

These apparently paradoxical associations are potentially attributable to changes in behaviour in response to the pandemic, whereby people at greater risk of severe disease because of older age or presence of comorbidities may reduce social contact and visits to indoor public places in order to reduce their exposure to SARS-CoV-2. However, to our knowledge, studies to investigate whether behaviours influencing risk of such exposure might partly explain associations between older age, presence of comorbidities and lower risk of developing COVID-19 are lacking. Such studies could potentially shed light on other controversies relating to risk factors for developing COVID-19, such as the extent to which ethnic differences in disease susceptibility can be explained by behavioural, occupational and socio-economic factors,^9^ and whether lifestyle, diet, and use of micronutrient supplements may influence risk of developing COVID-19.^10,11^

In order to address this knowledge gap, we established a new longitudinal study (COVIDENCE UK) at the start of the pandemic, with the specific aim of capturing detailed information on a very wide range of potential risk factors for COVID-19. Sufficient incident cases of test-confirmed COVID-19 have now accumulated to allow us to evaluate how a comprehensive panel of demographic, socio-economic, lifestyle, dietary, pharmacological and comorbidity factors relate to the risk of developing COVID-19.

## Methods

### Study design, setting and participants

COVIDENCE UK (Longitudinal population-based observational study of coronavirus disease in the UK population, www.qmul.ac.uk/covidence) is a prospective cohort study with four main objectives, namely to: i) determine risk factors for incident COVID-19 in the UK population; ii) characterise the natural history of COVID-19 in the UK population; iii) evaluate the impact of COVID-19 on the physical and mental health of the UK population; and iv) provide a resource from which to identify potential participants for future clinical trials of interventions to reduce incidence and/or severity of COVID-19 and other acute respiratory infections. Inclusion criteria were age 16 years or more and residence in the UK at the point of enrolment; there were no exclusion criteria. Participants were invited via a national media campaign to complete an on-line baseline questionnaire to capture information on potential symptoms of COVID-19 experienced since 1^st^ February 2020, results of any COVID-19 tests, and details of a wide range of potential risk factors for COVID-19, as described below. Follow-up questionnaires were administered at monthly intervals to capture incident test-confirmed COVID-19 as well as potential symptoms of COVID-19. The study was launched on 1^st^ May 2020, and this paper reports findings of analysis of data collected up to 5^th^ February 2021.

### Sponsorship, registration, ethics and reporting

The study was sponsored by Queen Mary University of London and approved by Leicester South Research Ethics Committee (ref 20/EM/0117). It is registered with ClinicalTrials.gov (NCT04330599).

### Outcomes

The primary outcome was incidence of test-confirmed COVID-19, as defined by a self-reported positive result from PCR or lateral flow testing of eluate from a nose or throat swab for SARS-CoV-2. Those who were not tested were assumed to be test negative. The secondary outcome was incidence of symptom-defined probable COVID-19, with casehood defined using the algorithm described by Menni and colleagues,^12^ based on age, sex and self-reported loss of smell/taste, significant/severe persistent cough, severe fatigue and skipped meals (see supplementary Appendix for further details). This outcome was included in order to address potential under-ascertainment of COVID-19 arising from use of test-confirmed COVID-19 as an outcome measure, which would not have captured episodes where testing was not done, potentially introducing collider bias.^13^ In order to minimise the potential for reverse causality to explain associations observed, outcomes occurring within 30 days of enrolment were censored. At enrolment, participants were asked to provide details of potential symptoms of COVID-19 experienced since 1^st^ February 2020, and results of any PCR or lateral flow tests for SARS-CoV-2 performed on eluates from nose / throat swabs to date.

### Independent variables

At enrolment, participants were asked to complete an on-line questionnaire capturing information about their socio-demographic characteristics, type of occupation, lifestyle, weight, height, longstanding medical conditions, medication use, vaccination status, diet and supplemental micronutrient intake (for baseline questionnaire, see Table S1, Supplementary Appendix). Monthly on-line follow-up questionnaires (Table S2, Supplementary Appendix) captured incident test-confirmed COVID-19 and potential symptoms of COVID-19.

### Sample size

The sample size required to detect an odds ratio of at least 1.08 (effect size) for a binary exposure variable with maximum variability (probability = 0.50 changing to 0.52) and correlated with other variables in the model (R^2^ = 0.5), with a power of 90% using a one-sided test with 5% significance level was estimated as 10,721, using the ‘powerlog’ program in Stata 14.2 (College Station, TX). Assuming 10% censoring at baseline (prevalent COVID-19 and missing) and 20% loss to follow-up, we aimed to recruit a minimum of 14,890 participants. No upper limit for sample size was specified.

### Statistical methods

Statistical analyses were performed using Stata 14.2. Putative risk factors for COVID-19 were selected *a priori* and classified into the following groups: socio-demographic, occupational and lifestyle factors; longstanding medical conditions, medication use and vaccination status; and diet and supplemental micronutrient intake. To produce patient-level covariates for each class of medications investigated, participant answers were mapped to drug classes listed on the British National Formulary (BNF) or the DrugBank and Electronic Medicines Compendium databases if not explicitly listed on the BNF; further details of the computational methods used to achieve this are presented in supplementary Appendix. Index of Multiple Deprivation (IMD) 2019 scores were assigned according to participants’ postcodes, and categorised into quartiles.

Participants who reported definite COVID-19 prior to enrolment, or who were classified as having had symptom-defined probable COVID-19 prior to enrolment on the basis of self-reported symptoms, were excluded from prospective analyses. Logistic regression models were used to estimate odds ratios (ORs) and 95% confidence intervals (CIs) for each individual factor, first in a crude model, then in ‘minimally adjusted’ models. For the primary outcome of test-confirmed COVID-19, the minimally adjusted model included age (six categories), sex (male/female), duration of participation and frequency of COVID-19 testing. For the secondary outcome of symptom-defined probable COVID-19, the minimally adjusted model included age (six categories), sex and duration of participation only. Factors associating independently with each outcome at the 10% significance level were evaluated for collinearity using Cramer’s V statistic^14^ for all pairwise combinations of the covariates and clustering with average-linkage hierarchical clustering using 1 - V as a dissimilarity metric. The resultant heat-maps were reviewed, and where a cluster of highly correlated covariates was identified, one variable within each cluster that was deemed to relate most closely and plausibly to COVID-19 risk was selected for inclusion in the multivariable model. Covariate associations with age were not deemed to warrant exclusion from the multivariable model, since all covariates investigated in the collinearity analysis exhibited a significant relationship with COVID-19 in the minimally-adjusted model that controlled for age. Variables that did not exhibit clustering in the heat-maps were all included in the final model, along with age, sex, IMD score quartile, duration of participation and frequency of COVID-19 testing. Correction for multiple comparisons was not applied, on the grounds that we were testing *a priori* hypotheses for all risk factors investigated.^15^ For the primary outcome, two sensitivity analyses were performed: one excluded participants who received one or more doses of COVID-19 vaccine, and the other excluded those who were randomised to receive vitamin D supplementation as part of a nested clinical trial that was initiated during follow-up.

## Results

Of the 17,558 participants who completed the COVIDENCE UK baseline questionnaire on or before 2^nd^ November 2020, we excluded those who were identified as already having had test-confirmed and/or symptom-defined probable COVID-19 (n=1,613) and those who did not complete at least one monthly follow-up questionnaire (n=854), leaving 15,227 participants eligible for inclusion in this prospective analysis (Figure S1, Supplementary Appendix). Selected baseline characteristics are presented in Table 1. Mean age was 59.4 years (range 16.0 to 94.4 years), 69.8% were female, and 94.9% identified their ethnic origin as White.

**Table 1.**
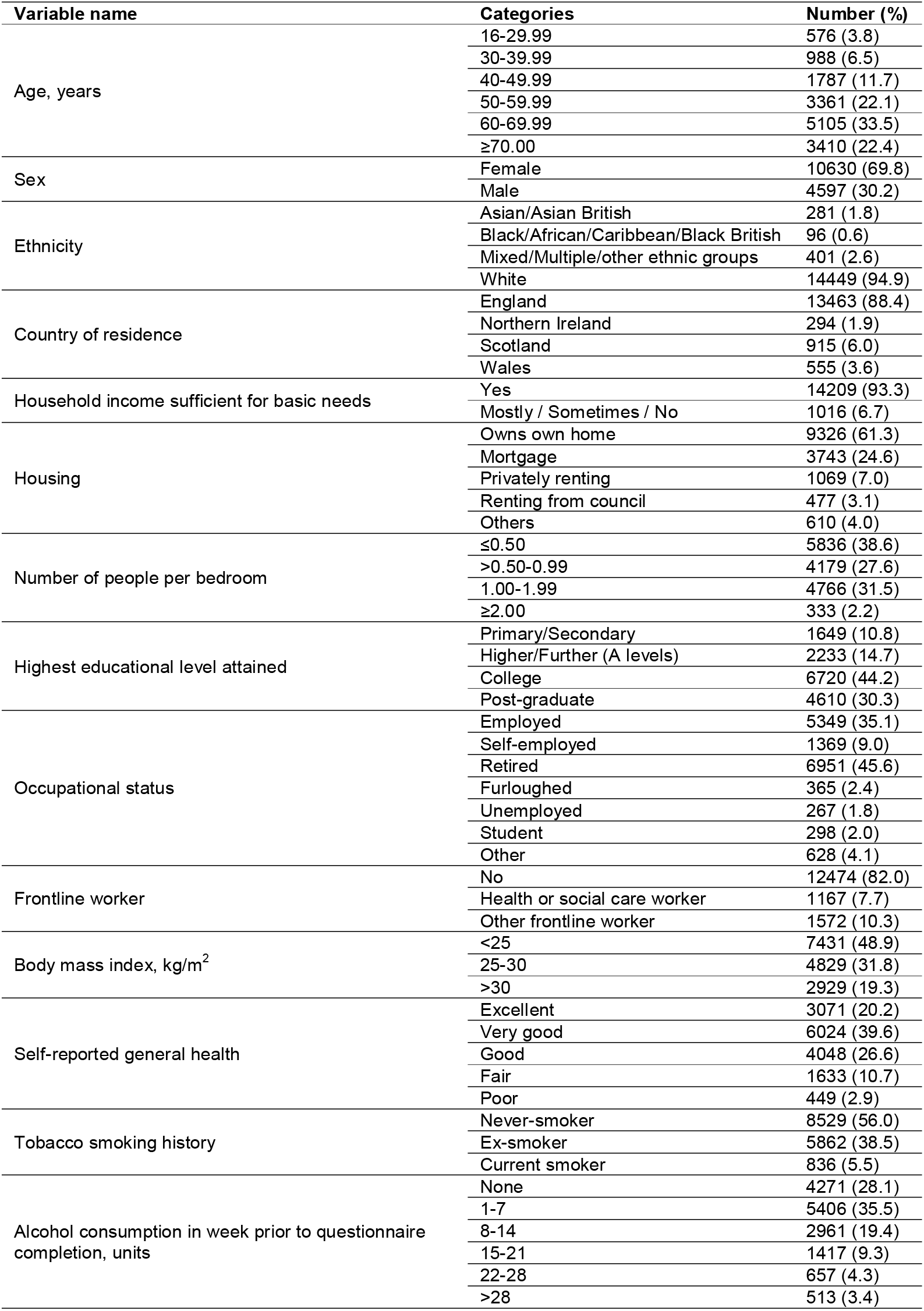
Selected cohort characteristics (n=15,227)

The geographical distribution of COVIDENCE UK participants aligned closely with that of incident COVID-19 in the UK (Figure S2, Supplementary Appendix).

A total of 446 participants experienced at least one episode of PCR- or lateral flow test-confirmed COVID-19 during 2,613,921 person-days of follow-up, of whom 32 were hospitalised. We calculated crude and minimally-adjusted ORs for associations between risk of test-confirmed COVID-19 and socio-demographic, occupational and lifestyle factors (Table 2); longstanding conditions, medication use and vaccination status (Table 3); and diet and supplemental micronutrient intake (Table 4). After adjustment for age, sex, duration of participation and testing frequency (‘minimal adjustment’), the following factors were found to associate with increased risk of COVID-19 with P <0.10: Asian / Asian British vs. White ethnic origin, housing type (paying mortgage and ‘other’ vs. owning own home), frontline vs. non-frontline worker status, household overcrowding (>0.5 vs. ≤0.5 people per bedroom), any visit vs. no visits to/from other households in the previous week, presence vs. absence of schoolchildren and working-age adults in the household, living with others vs. living alone, having vs. not having a dog at home, any vs. no travel to work or place of study in the week preceding questionnaire completion, number of visits to shops or other indoor public places per week (quartiles 2,3,4 vs quartile 1), alcohol consumption (15-21 vs 0 U/week); sleep duration (7 or ≥9 vs 8 hours per night); raised body mass index (BMI) (>25.0 vs ≤25.0 kg/m^2^) and history vs. no history of BCG vaccination. The following factors associated with decreased risk of COVID-19 with P<0.10 after minimal adjustment: age ≥60 vs. 16-29.99 years, education to college or postgraduate level vs primary / secondary level, shielding vs. non-shielding, low-impact physical activity (≥2 vs 0 hours/week), presence vs. absence of asthma diagnosis, presence vs. absence of atopic disease (defined by atopic eczema/dermatitis and/or hayfever/allergic rhinitis), use vs. no use of systemic immunosuppressants, inhaled corticosteroids and bronchodilators (defined as β-2-adrenoreceptor agonists or anticholinergics), higher intake of fruit and vegetables (top vs bottom quartiles) and use vs. no use of vitamin D supplements.

**Table 2.**
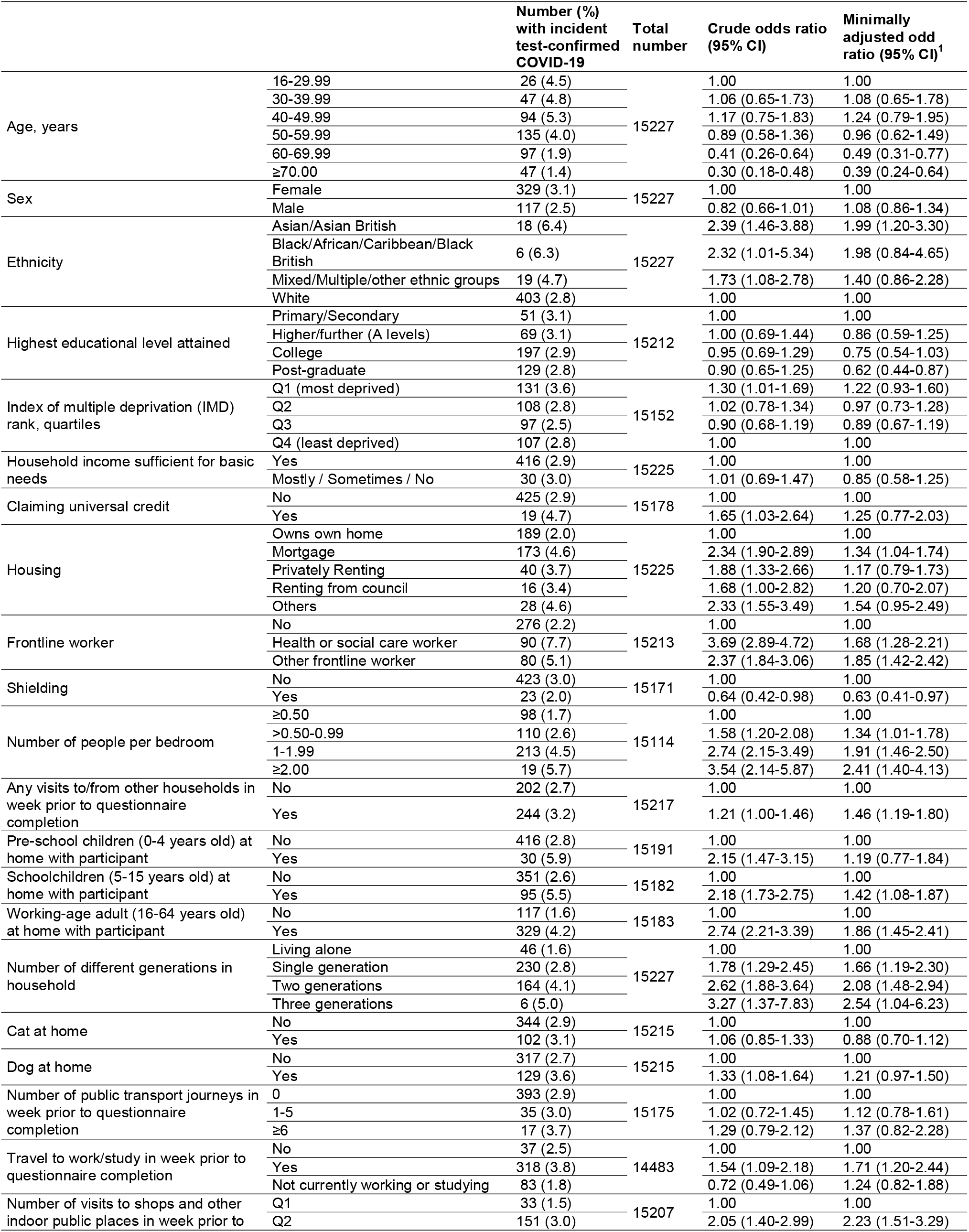
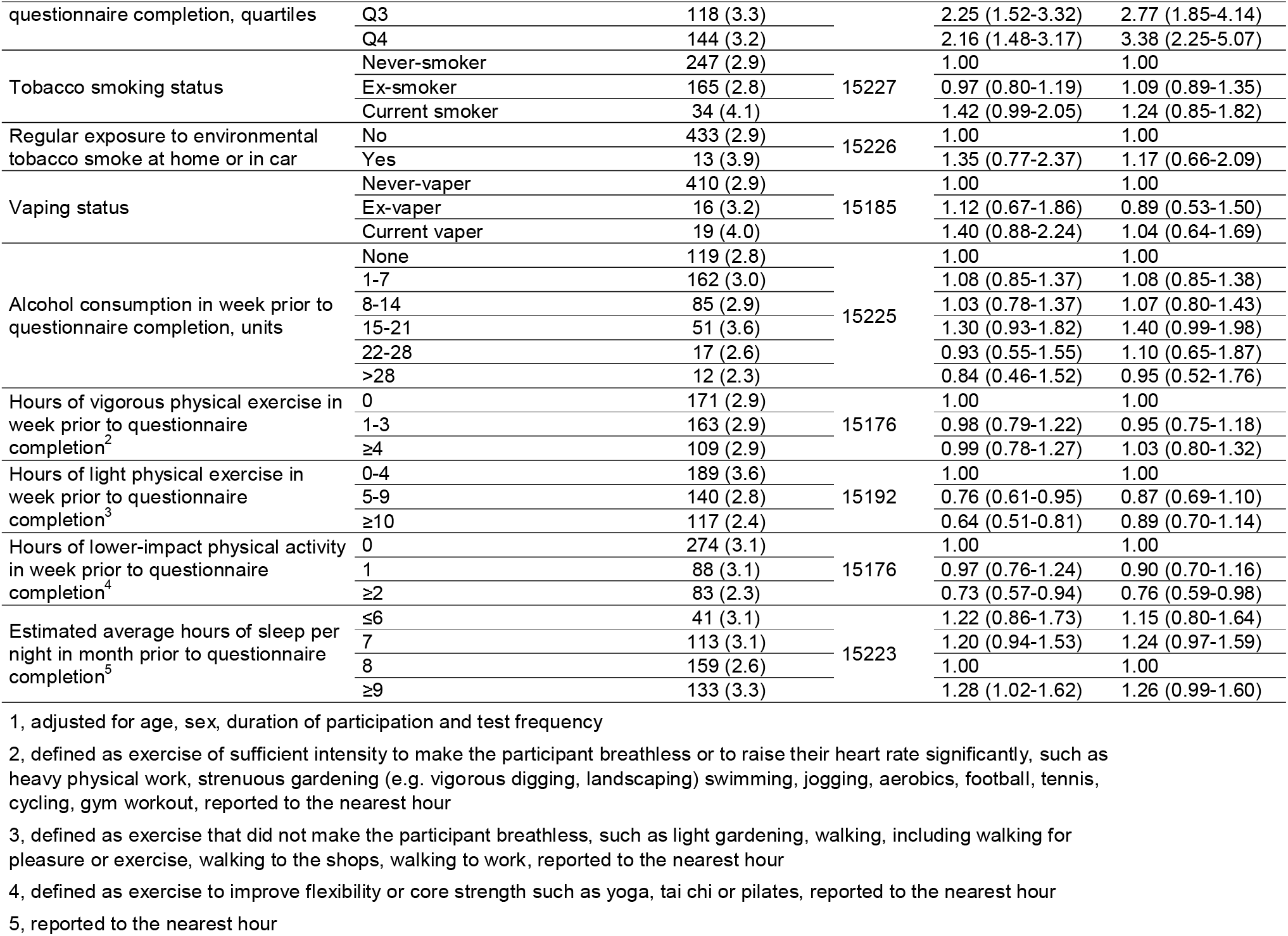
Socio-demographic, occupational and lifestyle factors and risk of test-confirmed COVID-19: crude and minimally-adjusted odds ratios

**Table 3.**
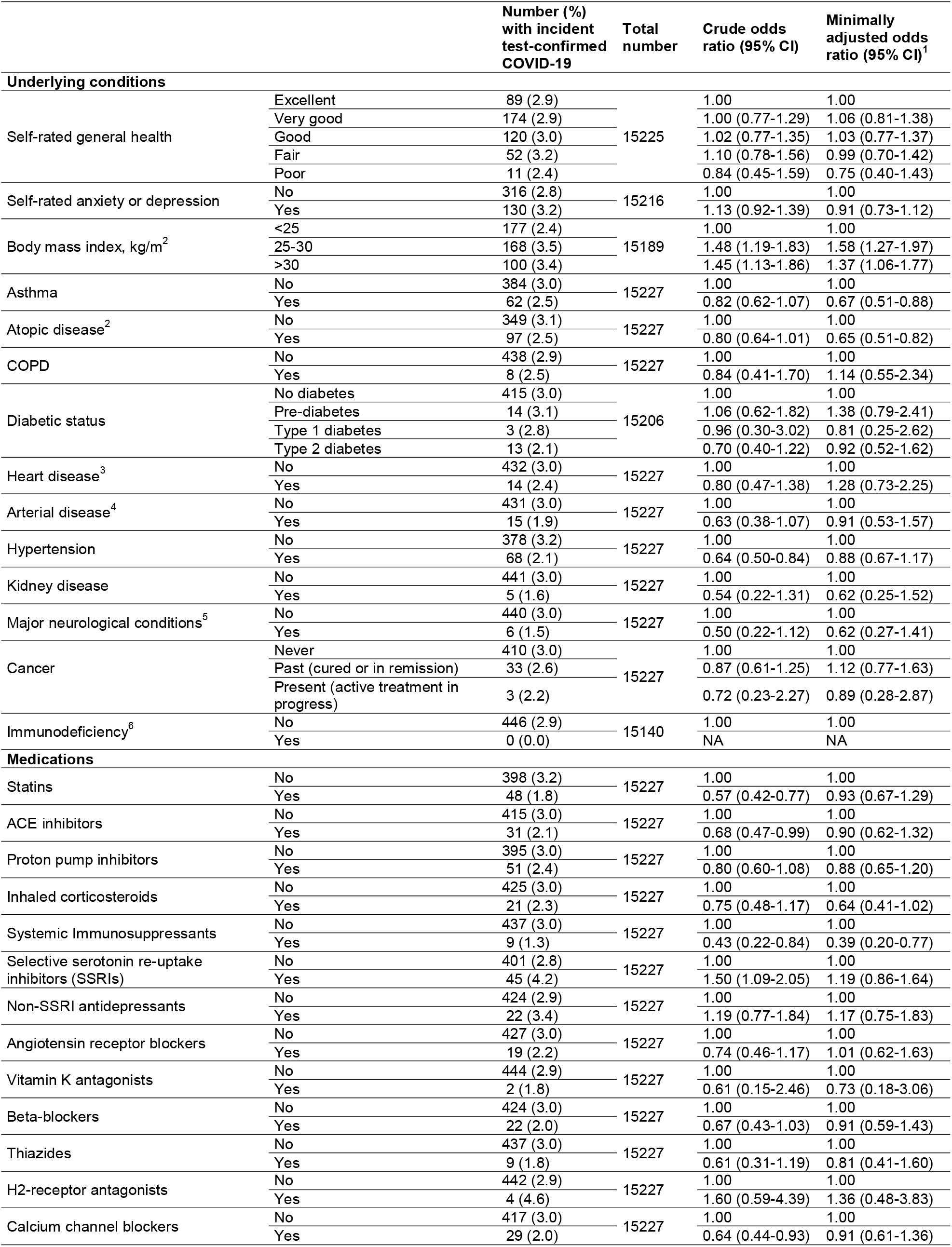
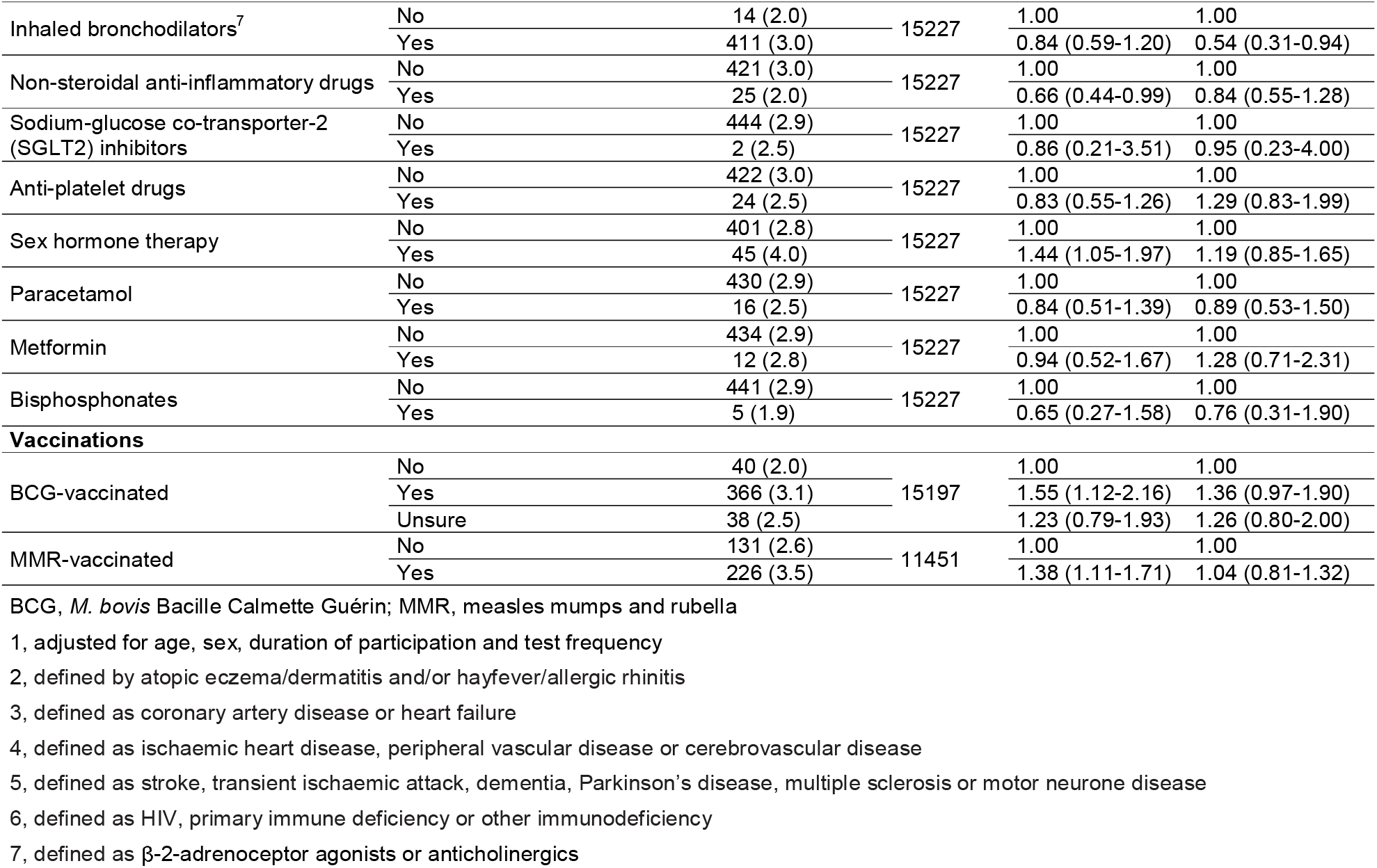
Underlying conditions, medication use, vaccination status and risk of test-confirmed COVID-19: crude and minimally-adjusted odds ratios

**Table 4.** Diet, supplemental micronutrient intake and risk of test-confirmed COVID-19: crude and minimally-adjusted odds ratios

|  |  | Number (%)<br>with incident<br>test-confirmed<br>COVID-19 | Total<br>number | Crude odds<br>ratio (95% CI) | Minimally<br>adjusted odds<br>ratio (95% CI)<sup>1</sup> |
| --- | --- | --- | --- | --- | --- |
| **Diet** |  |  |  |  |  |
| Dietary restrictions | None | 419 (2.9) | 15227 | 1.00 | 1.00 |
|  | Vegetarian<sup>2</sup> | 22 (3.2) |  | 1.11 (0.72-1.72) | 0.89 (0.57-1.40) |
|  | Vegan<sup>3</sup> | 5 (2.2) |  | 0.75 (0.31-1.83) | 0.60 (0.24-1.49) |
| Number of portions of fruit, vegetables, and salad intake per day in week prior to questionnaire completion, quartiles | Q1 | 78 (3.5) | 15188 | 1.00 | 1.00 |
|  | Q2 | 158 (3.1) |  | 0.88 (0.67-1.17) | 0.90 (0.68-1.20) |
|  | Q3 | 87 (3.0) |  | 0.85 (0.62-1.16) | 0.89 (0.64-1.22) |
|  | Q4 | 120 (2.4) |  | 0.66 (0.49-0.88) | 0.73 (0.54-0.99) |
| Number of portions of dairy products per day in week prior to questionnaire completion | 0-1 | 114 (2.8) | 15183 | 1.00 | 1.00 |
|  | 2 | 142 (3.2) |  | 1.15 (0.90-1.48) | 1.18 (0.91-1.53) |
|  | 3-5 | 99 (2.7) |  | 0.96 (0.73-1.27) | 1.06 (0.80-1.41) |
|  | ≥6 | 91 (2.9) |  | 1.03 (0.78-1.37) | 1.24 (0.93-1.65) |
| Total number of portions of fish (oily or white) intake in the week prior to questionnaire completion | Q1 | 94 (3.8) | 15213 | 1.00 | 1.00 |
|  | Q2 | 79 (3.2) |  | 0.85 (0.63-1.15) | 0.99 (0.73-1.36) |
|  | Q3 | 111 (2.7) |  | 0.71 (0.53-0.93) | 0.88 (0.66-1.18) |
|  | Q4 | 162 (2.6) |  | 0.70 (0.54-0.90) | 0.95 (0.72-1.25) |
| Portions of oily fish intake in the week prior to questionnaire completion | 0 | 210 (3.7) | 15220 | 1.00 | 1.00 |
|  | 1 | 133 (2.7) |  | 0.72 (0.58-0.90) | 0.86 (0.69-1.08) |
|  | 2 | 71 (2.4) |  | 0.64 (0.49-0.85) | 0.80 (0.60-1.06) |
|  | ≥3 | 32 (2.1) |  | 0.57 (0.39-0.83) | 0.76 (0.52-1.13) |
| Portions of white fish intake in the week prior to questionnaire completion | 0 | 131 (3.3) | 15216 | 1.00 | 1.00 |
|  | 1 | 170 (2.8) |  | 0.83 (0.66-1.04) | 0.99 (0.78-1.26) |
|  | 2 | 92 (2.6) |  | 0.76 (0.58-1.00) | 0.95 (0.72-1.26) |
|  | ≥3 | 53 (3.3) |  | 0.98 (0.71-1.35) | 1.16 (0.83-1.61) |
| Number of cups of non-alcoholic fluids per day in the week prior to questionnaire completion, quartiles | Q1 | 104 (3.0) | 15177 | 1.00 | 1.00 |
|  | Q2 | 75 (2.9) |  | 0.98 (0.73-1.32) | 1.03 (0.76-1.41) |
|  | Q3 | 116 (2.5) |  | 0.84 (0.64-1.10) | 0.81 (0.62-1.07) |
|  | Q4 | 149 (3.4) |  | 1.14 (0.88-1.47) | 0.99 (0.76-1.29) |
| **Micronutrient supplement use** |  |  |  |  |  |
| Multivitamin supplement | No | 331 (2.7) | 15227 | 1.00 | 1.00 |
|  | Yes | 115 (3.6) |  | 1.33 (1.07-1.65) | 1.12 (0.89-1.40) |
| Vitamin A supplement | No | 445 (2.9) | 15227 | 1.00 | 1.00 |
|  | Yes | 1 (1.1) |  | 0.38 (0.05-2.73) | 0.43 (0.06-3.15) |
| Vitamin C supplement | No | 397 (2.9) | 15227 | 1.00 | 1.00 |
|  | Yes | 49 (3.1) |  | 1.09 (0.80-1.47) | 1.03 (0.76-1.41) |
| Vitamin D supplement | No | 305 (3.2) | 15227 | 1.00 | 1.00 |
|  | Yes | 141 (2.5) |  | 0.80 (0.65-0.98) | 0.80 (0.65-0.99) |
| Zinc supplement | No | 425 (2.9) | 15227 | 1.00 | 1.00 |
|  | Yes | 21 (2.8) |  | 0.97 (0.62-1.51) | 0.93 (0.58-1.46) |
| Iron supplement | No | 423 (2.9) | 15227 | 1.00 | 1.00 |
|  | Yes | 23 (4.6) |  | 1.63 (1.06-2.51) | 1.27 (0.82-1.98) |
| Probiotic supplement | No | 426 (3.0) | 15227 | 1.00 | 1.00 |
|  | Yes | 20 (2.2) |  | 0.72 (0.46-1.13) | 0.69 (0.44-1.10) |
| Fish oil, krill oil or other omega-3 supplements | No | 400 (3.0) | 15227 | 1.00 | 1.00 |
|  | Yes | 46 (2.7) |  | 0.91 (0.67-1.25) | 0.98 (0.71-1.35) |
| Cod liver oil supplement | No | 409 (2.9) | 15227 | 1.00 | 1.00 |
|  | Yes | 37 (3.0) |  | 1.03 (0.73-1.45) | 1.29 (0.91-1.84) |
| Garlic or garlic powder (allicin) supplement | No | 439 (2.9) | 15227 | 1.00 | 1.00 |
|  | Yes | 7 (2.3) |  | 0.77 (0.36-1.63) | 0.93 (0.43-2.02) |
| Selenium supplement | No | 445 (3.0) | 15227 | 1.00 | 1.00 |
|  | Yes | 1 (0.6) |  | 0.21 (0.03-1.47) | 0.20 (0.03-1.46) |

1, adjusted for age, sex, duration of participation and test frequency
2, defined as excluding meat and fish, but not eggs or cow's milk, from the diet
3, defined as excluding meat, fish, eggs and cow's milk from the diet

All factors associating with test-confirmed COVID-19 with P <0.10 in the minimally-adjusted model were then assessed for collinearity: the resultant heat-map (Figure S3, Supplementary Appendix) revealed a high degree of collinearity between the number of working-age adults in the household, multi-generational households and number of people per bedroom. Since household over-crowding (as indicated by number of people per bedroom) was deemed to relate most closely to SARS-CoV-2 exposure risk, the other two independent variables were excluded from the multivariable model.

Table 5 presents fully-adjusted ORs for associations between potential risk factors for test-confirmed COVID-19. The final multivariable model adjusted mutually for age, sex, duration of participation, test frequency, ethnicity, highest educational level attained, IMD rank, household income, housing type, number of people per bedroom, presence of schoolchildren at home, presence of a dog in the household, shielding, visits to/from other households, visits to shops and other indoor places, travel to work or study, frontline worker status, low-impact physical activity, alcohol intake, BMI, history of asthma, history of atopic disease, use of systemic immunosuppressants, use of inhaled corticosteroids, use of bronchodilators, BCG vaccination status, intake of fruit, vegetables and salads, and intake of supplemental vitamin D. Increased risk of developing COVID-19 was independently associated with Asian/Asian British *vs*. White ethnicity (aOR 2.31, 95% CI 1.35-3.95), household overcrowding (aOR per additional 0.5 people/bedroom 1.26, 1.11-1.43), any *vs*. no visits to/from other households in previous week (aOR 1.33, 1.07-1.64), number of visits to indoor public places (aOR per extra visit per week 1.05, 1.01-1.09), frontline occupation outside health/social care *vs*. no frontline occupation (aOR 1.49, 1.12-1.98), and raised BMI (aOR 1.51 [1.20-1.90] for BMI 25.0-30.0 kg/m^2^ and 1.38 [1.05-1.82] for BMI >30.0 kg/m^2^ *vs*. BMI <25.0 kg/m^2^). Lower risk of test-confirmed COVID-19 was independently associated with history of atopic disease (aOR 0.76, 0.59-0.98) and taking systemic immunosuppressants (aOR 0.43, 0.19-0.95).

**Table 5.**
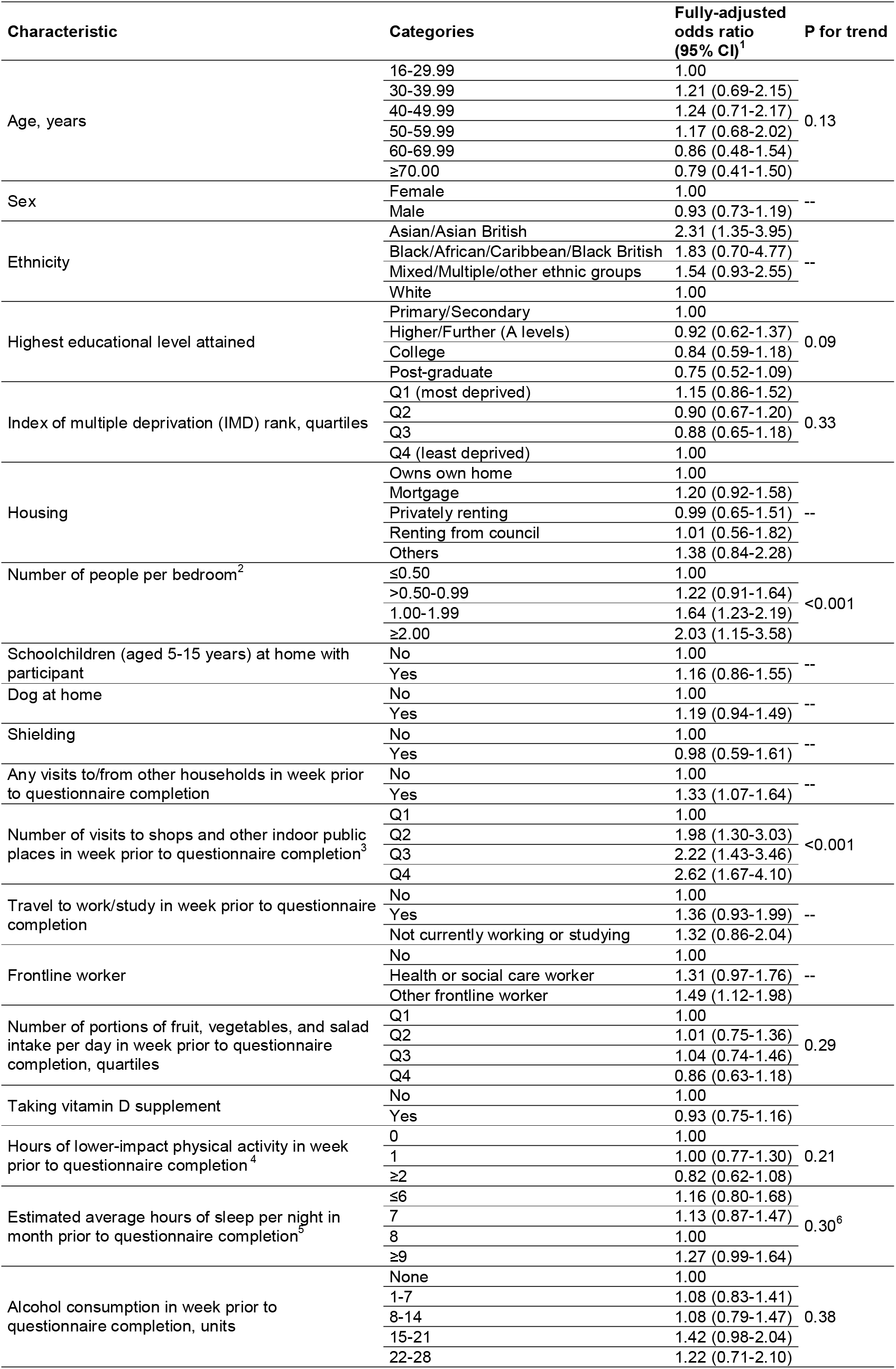
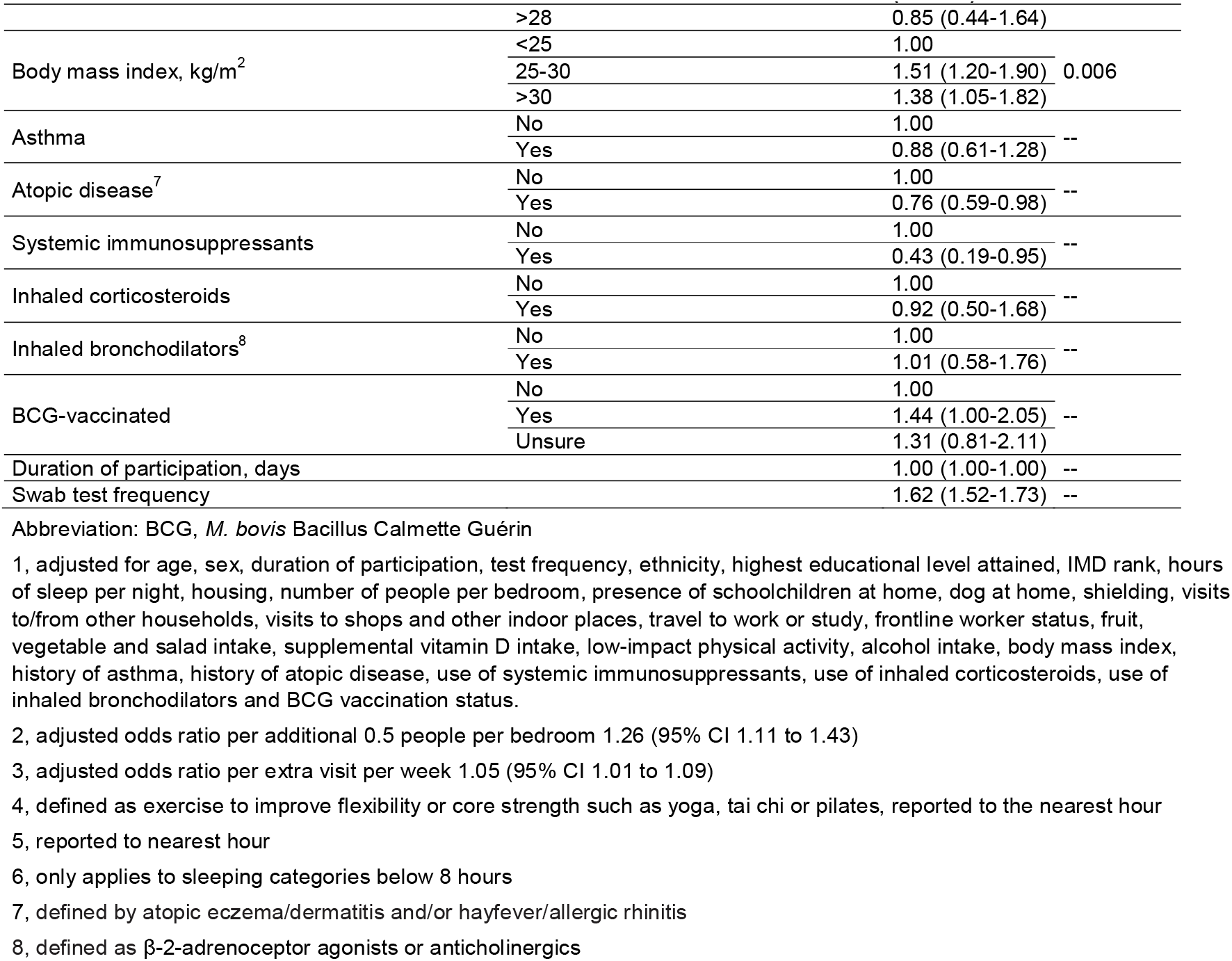
Independent risk factors for test-confirmed COVID-19: final multivariable model (n=14,027)

We performed two sensitivity analyses to explore the robustness of the multivariable results. The first (Table S3, Supplementary Appendix) excluded 3202 participants who received one or more doses of COVID-19 vaccine before the date of the data download (5^th^ February 2021). The second (Table S4, Supplementary Appendix) excluded 3813 participants who commenced vitamin D supplements after enrolment in the cohort due to participation in a clinical trial. Both analyses yielded similar findings to those presented above.

We also performed an exploratory analysis to determine whether COVID-19 risk differed for participants with atopic vs. non-atopic asthma endotypes, as defined by the presence or absence of ‘atopic disease’ (atopic eczema/dermatitis and/or hayfever/allergic rhinitis). This showed a reduced risk of COVID-19 for participants with atopic asthma (aOR 0.64, 0.42-0.96), but not for those with non-atopic asthma (aOR 0.91, 0.62-1.33), as compared to participants without atopic disease or asthma. These effect estimates did not materially change after further adjustment for inhaled corticosteroids (Table S5, Supplementary Appendix).

We then proceeded to investigate determinants of symptom-defined probable COVID-19, with casehood ascribed using an algorithm published by Menni et al.^12^ In the subset of COVIDENCE UK participants who had one or more tests for COVID-19, this case definition had sensitivity and specificity for test-confirmed COVID-19 of 0.68 and 0.78, respectively, with an area under the receiver operating characteristic curve of 0.79 (95% CI, 0.76 to 0.82). Potential risk factors associating with probable symptom-defined COVID-19 with P<0.10 in a minimally-adjusted model (i.e. adjusting for age, sex and duration of participation) were included in the multivariable model presented in Table S6, Supplementary Appendix. Increased risk of probable symptom-defined COVID-19 was independently associated with Asian / Asian British vs. White ethnicity, housing type (having a mortgage vs. home-ownership), household overcrowding (>0.5 vs. ≤0.5 people per bedroom), frontline occupation outside of health or social care, use of cod liver oil supplements, poorer self-assessed general health, and use of selective serotonin reuptake inhibitors, while lower risk of probable symptom-defined COVID-19 was independently associated with greater age (age ≥50 vs. 16-29.99 years).

## Discussion

In this large, prospective population-based study evaluating a diverse array of potential risk factors for developing COVID-19, we found that Asian/Asian British ethnicity, household overcrowding, indoor social mixing, employment as a frontline worker outside of health and social care, and being overweight or obese were all independently associated with an increased risk of test-confirmed COVID-19.

Associations with household overcrowding and visits to indoor public places showed dose-response relationships, supporting a causal interpretation. History of atopic disease and use of systemic immunosuppressant medication were independently associated with decreased risk of test-positive disease. No statistically significant independent associations with disease risk were seen for other factors investigated, including age, sex, diet, supplemental micronutrient intake, and other longstanding conditions and medications.

This study sheds new light on the degree of overlap between risk factors for developing COVID-19 (irrespective of severity) *vs*. risk factors for developing severe or fatal disease specifically. Our finding that people of Asian/Asian British ethnic origin are at increased risk of developing COVID-19 is consistent with reports of increased susceptibility and disease severity in this group.^4,7,16^ One limitation of previous studies investigating ethnic variation in COVID-19 risk is that they did not adjust for behaviours influencing SARS-CoV-2 exposure, such as visits to other households and indoor public places. In our study, increased risk of developing COVID-19 in people of Asian/Asian British ethnic origin was not explained by such behaviours, nor by social deprivation, domestic overcrowding, occupation, BMI, or comorbidities. There is therefore an urgent need to explain ethnic disparities in risk of developing COVID-19 so that preventive strategies can be deployed. The association between raised BMI and increased susceptibility to COVID-19 that we found is consistent with studies identifying obesity as a risk factor for both susceptibility to, and severe outcomes of, COVID-19.^4,17,18^ It would appear that immune dysregulation associated with obesity may increase susceptibility to infection as well as disease severity.

By contrast, a number of established risk factors for severe and fatal disease, including older age, male sex and underlying conditions such as diabetes, heart disease, COPD and hypertension, were not associated with risk of developing COVID-19 in our study. In keeping with reports from the UK^7^ and elsewhere,^8^ we found younger age to be associated with increased risk of developing COVID-19 in crude and minimally-adjusted models. However, this association did not persist after adjustment for multiple potential confounders, including behaviours related to social mixing, suggesting that lower incidence of COVID-19 in older adults in our study may be explained by reductions in social contact. We did not see a difference in disease risk for people with diabetes, heart disease or hypertension. Whilst this contrasts with a study reporting lower prevalence of SARS-CoV-2 seropositivity among people with these underlying conditions, that study did not adjust, as we did, for behaviours influencing exposure to infection.^7^ The only long-standing conditions associated with disease risk in our study were atopic diseases, which were associated with reduced risk of disease, particularly among those who also had asthma. This may reflect decreased expression of *ACE2*, the SARS-CoV-2 receptor, which has been reported in people with both high levels of allergic sensitisation and asthma.^19^

Our study has several strengths. COVIDENCE UK was set up with the specific purpose of investigating incident COVID-19, and consequently our questionnaires were specifically designed to capture contemporaneous and granular detail on potential risk factors, including behaviours influencing risk of exposure to SARS-CoV-2. Our finding that visits to other households and indoor public places were associated with increased risk of disease supports the case for restricting such activities as a public health strategy to control disease. Our low rates of loss to follow-up reflect the very high degree of participant engagement with the COVIDENCE UK study. Our ability to identify episodes of milder disease affords potential insights into susceptibility factors as well as severity factors, and sets our study apart from long-established cohort studies in which assessment of risk factors may be temporally remote, and capture of outcomes is limited to events that are fatal or that precipitate hospitalisation. Our prospective design, coupled with censoring events occurring within 30 days of enrolment minimises the potential for reverse causation to explain associations observed.

Our study also has limitations. Use of test-confirmed COVID-19 as our primary outcome may have resulted in under-ascertainment of disease, particularly early in the pandemic (when testing capacity was particularly limited) and among people with less access to testing services; this might introduce collider bias.^13^ We addressed this limitation by including a secondary outcome of symptom-defined probable COVID-19, which did not rely on access to testing. A second issue relates to under-representation of ethnic minorities, particularly people of Black, African and Caribbean ethnic origin; a lack of statistical power may explain why we did not confirm an increased risk of disease in these groups. Third, as with any observational study, we cannot exclude the possibility that the associations we report may be explained by residual and/or unmeasured confounding. We have sought to minimise this by comprehensive capture of, and rigorous adjustment for, multiple potential confounders of relationships between putative risk factors and our primary outcome.

In conclusion, this population-based longitudinal study conducted in UK adults found that increased risk of developing COVID-19 associated independently with Asian/Asian British ethnicity, household overcrowding, visits to other households and other indoor public places, frontline occupation outside of health or social care, and increased BMI, after rigorous adjustment for multiple confounders. Atopic diseases, and especially atopic asthma, were associated with decreased risk. In contrast to studies investigating risk factors for severe disease, older age, male sex and other comorbidities were not associated with increased risk of developing COVID-19.

## Supporting information

Supplementary Appendix Holt et al

## Data Availability

A copy of anonymised data will be made available following publication of this manuscript in a peer-reviewed journal

## Acknowledgements

This study was supported by a grant from Barts Charity to ARM and CJG (ref MGU0466). The work was carried out with the support of BREATHE - The Health Data Research Hub for Respiratory Health [MC_PC_19004] in partnership with SAIL Databank. BREATHE is funded through the UK Research and Innovation Industrial Strategy Challenge Fund and delivered through Health Data Research UK. MT is supported by a grant from the Rosetrees Trust and The Bloom Foundation (ref: M771). The views expressed are those of the authors and not necessarily those of Barts Charity, BREATHE or Health Data Research UK. We thank all the people who participated in the COVIDENCE UK study, and the following individuals and organisations who supported study recruitment: Asthma UK, the British Heart Foundation, the British Lung Foundation, the British Obesity Society, Cancer Research UK, Diabetes UK, Future Publishing Ltd, Kidney Care UK, Kidney Wales, Mumsnet, the National Kidney Federation, the National Rheumatoid Arthritis Society, the North West London Health Research Register (DISCOVER), Primary Immunodeficiency UK, the Race Equality Foundation, SWM Health Ltd, the Terence Higgins Trust and Vasculitis UK.

## Author Contributions

ARM wrote the study protocol, with input from HH, MT, CR, GB and SOS. HH, MT, JS, CR, KY, MD, KT, SF, SI, AM, PP, DM, GD, RL, CJG, FK, AS, GB, SAS and ARM contributed to questionnaire development and design. HH co-ordinated and managed the study, with input from ARM, MT, JS and SOS. HH, JS, ARM, SOS, NSH and BA supported recruitment. HH, MT, MG, MD, KT, SSR, AAK, SER, PJL and DAJ contributed to data management and coding medication data. Statistical analyses were done by MT, with input from SOS, ARM, MG and HH. ARM wrote the first draft of the report. All authors revised it critically for important intellectual content, gave final approval of the version to be published, and agreed to be accountable for all aspects of the work in ensuring that questions related to the accuracy or integrity of any part of the work were appropriately investigated and resolved.

## Transparency Declaration

ARM is the manuscript’s guarantor and he affirms that this is an honest, accurate, and transparent account of the study.

