## Supplementary Appendix Holt et al for "Risk factors for developing COVID-19: a population-based longitudinal study (COVIDENCE UK)"

### **Supplementary Methods**

#### *Symptom-based algorithm*

We used the following equation previously described and validated by Menni and colleagues^1^ to assign probable COVID-19 status:

Predicted COVID-19 = -(0.01 x age) + (0.44 x sex) + (1.75 x loss of smell/taste) + (0.31 x persistent cough) + (0.49 x severe fatigue) + (0.39 x skipped meals).

All symptoms were coded as 1 if the participant reported the symptom, and 0 if not. Male vs female sex were coded as 1 vs 0, respectively. The obtained value was then transformed into a predicted probability of COVID-19 using exp(*x*)/(1 + exp(*x*)) transformation. Presence of probable COVID-19 was assigned for predicted probabilities >0.5.

#### *Medication classification*

The raw survey answers describing the names of medications prescribed to each participant were first processed using regular expressions to remove text related to dosage, route of administration, formulation, and frequency of administration. Each participant’s set of processed medication answers were then mapped to a curated, composite database containing 135,167 drug aliases (and their active ingredients) sourced from both the DrugBank and Electronic Medicines Compendium (EMC) databases. This mapping consisted of four steps. First, the composite database was searched for exact matches with each of the processed survey answers. For survey answers with no exact matches, the database was searched for an exact match with the first word of each survey answer. Then, the Metaphone algorithm ^2^ was applied to produce a phonetic encoding for each of the remaining unmapped survey answers and the entire composite drug database. Survey answers with an unambiguous, exact phonetic match with an alias in the database were annotated accordingly. Finally, for survey answers that could not be mapped in any of the three steps detailed above, Levenshtein distance (LD) ^3^ values were calculated between each unmapped survey answer and every alias in the drug database. For each processed survey answer, drug aliases in the database with an LD of 1 from the answer were identified. If the survey answer mapped to a single drug alias in the database with an LD of 1, the answer was annotated accordingly. If the survey answer mapped to multiple entries in the drug database with an LD of 1, the drug alias with the active ingredients appearing at the highest mean frequency across the entire annotated data set was selected. The remaining survey answers - for which no aliases in the database returned an LD value of 1 - were annotated manually.

With the active ingredient mapping for each survey answer, participant-level annotations were produced by considering each participant’s total set of survey answers. One set of covariates was produced by annotating each participant with a value of 1 for drug classes containing one or more compounds included in their survey answers and a value of 0 for all other drug classes. Another set of covariates was produced by annotating each participant with a scaled dosage value for each drug class, again taking a value of 0 for classes for which the participant did not report any medications. Z-score normalisation of dosages was applied separately for each active ingredient in each class. The dosage z-scores were subsequently transformed using a probit function, normalising them to values in the interval (0, 1) and allowing comparison between participants not taking any drugs from the class (dosage = 0) and participants taking varying dosages of drugs from the class (0 < dosage < 1). Code is available at [https://github.com/mgreenig/COVIDENCE-survey](https://eur01.safelinks.protection.outlook.com/?url=https%3A%2F%2Fgithub.com%2Fmgreenig%2FCOVIDENCE-survey&data=04%7C01%7C%7C62d495b62f724102264c08d8a423d2ca%7C569df091b01340e386eebd9cb9e25814%7C0%7C0%7C637439821203279159%7CUnknown%7CTWFpbGZsb3d8eyJWIjoiMC4wLjAwMDAiLCJQIjoiV2luMzIiLCJBTiI6Ik1haWwiLCJXVCI6Mn0%3D%7C1000&sdata=QWiOn%2Bl1kySG53HIdt1dy0rdOjNmDk0Et%2FEDlnTNMpA%3D&reserved=0)

### **Supplementary Tables**

#### **Table S1.** Baseline questions

| **Sociodemographic** | |
| --- | --- |
| Date of birth (DD/MM/YYYY) |  |
| Post code |  |
| Address |  |
| Please state your **assigned sex at birth.** | -Male  -Female |
| What is your ethnic origin? | - White   - English / Welsh / Scottish / Northern Irish / British - Irish - Gypsy or Irish Traveller - Any other white background   - Mixed / Multiple ethnic groups   - White and Black Caribbean - White and Black African - White and Asian - Any other Mixed / Multiple ethnic backgrounds   - Asian / Asian British   - Indian - Pakistani - Bangladeshi - Chinese - Any other Asian background   - Black / African / Caribbean / Black British   - African - Caribbean - Any other Black / African / Caribbean background   - Arab  - Other Ethnic Group |
| Were you born in the UK? | - Yes  - No |
| Are you a ‘frontline worker’ who has to physically travel to work during lockdown?    Examples include people employed in health and social care, education and childcare, local/national government, food production or sale, the prison service, the police and public transport. | - Yes  - No |
| What is the highest level of education that you have completed? | - Primary school (0)  - Secondary school up to 16 years (1)  - Higher or secondary or further education (A-levels, BTEC, etc.) (2)  - College or university (3)  - Post-graduate degree (4) |
| In the last month, was your household income sufficient to cover the basic needs of your household, such as food and heating? | - Yes  - Mostly  - Sometimes  - No |
| Please select the box that best describes your current housing situation: | - I own my home outright  - I own my home and I am paying a mortgage  - I am renting privately  - I am renting from the council/housing association  - I am staying with friends or family  - I am homeless or living in temporary accommodation  - Other |
| Do you currently claim Universal Credit? | - Yes, I have applied to receive Universal Credit but have **not yet** received any payments  - Yes, I have claimed Universal Credit and received **one or more** payments  - No |
| How many bedrooms are there in your current accommodation? | - 1  - 2  - 3  - 4  - 5  - 6  - 7  - 8  - 9  - 10 + |
| Do you live alone? | - Yes  - No |
| How many people other than yourself live in your household? | - Children aged 0-4 years   - 0-10+   - Children aged 5-15 years   - 0-10+   - People aged 16-64 years   - 0-10+   - People aged 65 years or more   - 0-10+ |
| Does your household have any pets? | - Yes  - No |
| Which of the following best describes your current occupational status? | - Employed  - Self-employed  - Retired  - Furloughed  - Unemployed  - Student  - Other |
| Please indicate which types of pet you have at home.  Select all that apply | - Cat  - Dog  - Indoor bird (e.g. budgie, parrot, canary)  - Rabbit / Guinea pig / Hamster  - Tortoise, turtle, lizard or snake  - Other |
| **Comorbidities** | |
| What is your **current** height?  (if you are unsure, please put your best estimate) | -Feet/inches  -Centimetres |
| What is your **current** weight? | - Stones (sts) / pounds (lbs)  - Kilograms (kg) |
| Have you ever been diagnosed with any of the following conditions by a doctor? Select all that apply | - Asthma  - Atopic Eczema or Atopic Dermatitis  - Autoimmune disease (e.g. rheumatoid arthritis, multiple sclerosis (MS), lupus (SLE), Crohn’s disease,  ulcerative colitis, psoriasis, Raynaud’s disease, scleroderma)  - Cancer  - Cerebral Palsy  - COPD (including chronic bronchitis, and emphysema)  - Cystic Fibrosis  - Dementia  - Diabetes or pre-diabetes  - Hayfever or Allergic Rhinitis  - Heart Attack, Angina or Coronary Artery Disease  - Heart Failure  - High Blood Pressure (Hypertension)  - HIV Infection  - Hyperparathyroidism (overactive parathyroid gland)  - Kidney stones  - Other kidney disease  - Leg Artery Disease (also known as ‘peripheral vascular disease’, ‘peripheral arterial disease’ or ‘intermittent claudication’)  - Mental health disorder  - Motor Neurone Disease  - Organ transplant  - Parkinson's Disease  - Primary immune deficiency (e.g. antibody deficiency, combined immunodeficiency)  - Sarcoidosis  - Sickle Cell Disease (i.e. two copies of altered gene, affected by anaemia and other complications  - Sickle Cell Carrier (also known as ‘sickle cell trait’, with only one copy of altered gene: few symptoms if any)  - Splenectomy (removal of spleen)  - Stroke or Mini-Stroke  - Tuberculosis (TB)  - None of the above |
| You indicated you have been diagnosed with diabetes or pre-diabetes. Please specify your diagnosis: | - Pre-diabetes (high blood sugar levels, not enough to be diagnosed with diabetes)  - Type 1 diabetes  - Type 2 diabetes  - Other type of diabetes |
| Do you currently have cancer? | - Never  - No, cancer cured or in remission  - Yes, currently receiving treatment |
| Under each heading, please click the ONE box that best describes your health TODAY.  Anxiety / Depression | - I am not anxious or depressed  - I am moderately anxious or depressed  - I am extremely anxious or depressed |
| Over the last 12 months, would you say that on the whole, your health has been: | - Excellent  - Very good  - Good  - Fair  - Poor |
| **Vaccination** | |
| Have you ever had the BCG vaccine?   *This is the vaccine against Tuberculosis (TB), it's injected in the upper arm and usually leaves a small scar* | - Yes  - No  - Unsure |
| Have you ever had the MMR vaccine? *This is the vaccine against measles, mumps and rubella.*   *This is the vaccine against measles, mumps and rubella.* | - Yes  - No  - Unsure |
| **Lifestyle** | |
| Which of these best describes your use of cigarettes? | - I have never smoked cigarettes  - I used to smoke cigarettes occasionally but now not at all  - I used to smoke cigarettes daily but now not at all  - I smoke cigarettes occasionally but not every day  - I smoke cigarettes daily |
| Which of these best describes your use of e-cigarettes (vaping)? | - I have never vaped or used e-cigarettes  - I used to use e-cigarettes occasionally, but now not at all  - I used to use e-cigarettes daily but now not at all  - I vape occasionally but not every day  - I vape daily |
| Are you regularly exposed to smoke from other people’s cigarettes at home or in a car? | - Yes  - No |
| During the last week, roughly how many hours did you spend doing more vigorous physical exercise of sufficient intensity to make you breathless or to raise your heart rate significantly, such as heavy physical work, more strenuous gardening (e.g. vigorous digging, landscaping) swimming, jogging, aerobics, football, tennis, cycling, gym workout? | - 0 -10+ hours |
| During the last week, roughly how many hours did you spend doing lower impact physical exercise to improve flexibility or core strength such as yoga, tai chi or pilates? | - 0 -10+ hours |
| During the last week, roughly how many hours did you spend doing light exercise that does not make you particularly breathless, such as light gardening, walking, including walking for pleasure or exercise, walking to the shops, walking to work? | - 0 -10+ hours |
| During the past month, how many hours of actual sleep did you get per night on average?  (This may be different than the number of hours you spent in bed) | - 0 -24 hours |
| **Diet** | |
| Do you **exclude** any of the following foods from your diet? Select all that apply. | - Eggs  - Cow’s milk or products made from cow’s milk (e.g. cheese, yoghurt)  - Fish  - White meat (e.g. poultry)  - Red meat  - No, I eat all of these foods |
| Over the **last week**, how many **portions** of the following did you eat **per day**, on average?  Fruit, vegetables and salad?    *1 portion = 80g (e.g. one apple or two broccoli spears or 3 tablespoons of peas or carrots or one bowl of salad)* | - 0 – 10 or more |
| Over the **last week**, how many **portions** of the following did you eat **per day**, on average?  Dairy products (e.g. cow’s milk, cheese, yoghurts) or calcium-fortified dairy alternatives (e.g. soya milks, soya yoghurts and soya cheeses)    *1 portion = a cup of milk, a standard pot of yogurt or a piece of cheese about the size of two thumbs together (30g).* | - 0 – 10 or more |
| Over the last week, how many portions of **oily** fish did you eat?  e.g. herring, pilchards, salmon, sardines, sprats, trout and mackerel.    *1 portion = a small tin of oily fish (around 100g) or a piece of oily fish about the size of your palm* | - 0 – 10 or more |
| Over the last week, how many portions of **white fish or seafood (i.e non-oily fish)** did you eat?  e.g. cod, haddock, plaice, prawns and tuna  *1 portion = a small (160g) tin of tuna or a piece of white fish about the size of your palm (140g) or 100g prawns* | - 0 – 10 or more |
| Over the last week, how many cups or glasses of fluid did you drink per day, on average?   *1 cup or glass = about 150mL. Non-alcoholic drinks including water, tea, coffee, milk and other soft drinks all count* | - 0 -10+ |
| How many units of alcohol did you drink over the last 7 days?    One unit is a ½ a pint (285 ml) of ordinary beer, lager or cider; 25ml of spirits; 1 small glass (75ml) of wine; or 50ml of sherry. | - None  - 1-7 units  - 8-14 units  - 15-21 units  - 22-28 units  - More than 28 units |
| Over the **last month**, have you taken any of the following supplements at least **once per week**?  Select all that apply. | - Multivitamin (including prenatal multivitamins)  - Supplement containing vitamin A only  - Supplement containing vitamin B only  - Supplement containing vitamin C only  - Supplement containing vitamin D only  - Supplement containing calcium only  - Supplement containing calcium and vitamin D combined  - Supplement containing vitamin E only  - Supplement containing zinc only  - Supplement containing iron only  - Supplement containing probiotics  - Supplement containing fish oil, krill oil or other source of omega-3 fatty acids  - Supplement containing cod liver oil  - Supplement containing echinacea  - Supplement containing garlic or garlic powder (allicin)  - Supplement containing turmeric / curcumin  - Supplement containing Cannabidiol (CBD) oil  - Supplements containing folic acid  - Supplement containing Selenium only  - Other (e.g. other micronutrients (such as herbal supplements) or combinations of micronutrients (such vitamin C & zinc)) Please specify:  - None of the above |
| **Medications** | |
| Please type the names of all the medications you are currently taking below, one medication per box. The next pages will collect details about dosage for each.   Include all types of medications taken at home or administered in a hospital or clinic (capsules, tablets, contraceptive pills or implants, inhalers, injections, intravenous infusions, monoclonal antibodies, chemotherapy, immunosuppressants, etc.)   Please note that other pages will collect details about the amount of medicine in each dose (next page) and how often you take each dose (the page after that). If there are any details about your medication that aren’t captured by our form (e.g. if you take different doses of a medicine at different times of day), there will be space to enter them in a blank text box at the end of this section of the questionnaire. | 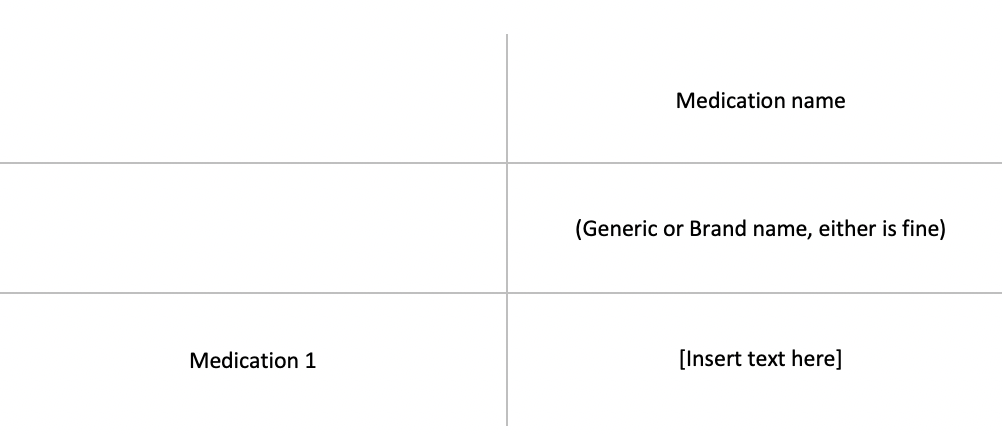 |
| Please select the frequency and route that you take your medication: | - Frequency   - 4 times/day - 3 times/day - 2 times/day - Once daily - Weekly - Less often than weekly - As needed   - Route   - By mouth - Inhaled - Injected - Other |
| **Recent health** | |
| Since February 1st 2020, have you experienced any of the following symptoms: loss of smell or taste, fever, persistent cough, fatigue, diarrhoea, abdominal pain or loss of appetite? | - Yes, I have had one or more of these symptoms since 1st of February  - No, I have not had any of these symptoms since 1st of February |
| When did your symptoms start? (DD/MM/YYYY) *e.g. 25/04/2020* |  |
| Did you have a persistent cough (coughing a lot for more than an hour, or 3 or more coughing episodes in 24 hours)? | - No  - Persistent dry cough (i.e. producing little or no phlegm)  - Persistent productive cough |
| Did you experience unusual fatigue? | - No  - Mild fatigue  - Severe fatigue - I struggled to get out of bed |
| Did you have a loss of sense of smell or taste? | - Yes  - No |
| Did you skip any meals because you felt unwell? | - Yes  - No |
| Since February 1st 2020, have you had a nose/throat swab to test for COVID-19? | - Yes  - No |
| On what date did you have this nose/throat swab?  If you are not sure of the exact date, enter the approximate date (DD/MM/YYYY).  *e.g. 25/04/2020* |  |
| What was the result? | - Positive  - Negative  - Not known |

#### **Table S2.** Follow-up questions

| **Since you last checked in with us**, have you had a nose or throat swab for COVID-19 or any other respiratory virus, or has a result from a previous swab test become newly available?(This question is about tests to detect the virus itself: they are usually done in somebody who has symptoms, but screening of asymptomatic people can also be done. It’s usually a nose/throat swab, but saliva tests are also becoming available) | - Yes  - No |
| --- | --- |
| On what date did you have this nose / throat swab? If you are not sure of the exact date, enter the approximate date (DD/MM/YYYY). |  |
| What was the result? Click as many as apply. | - Positive for COVID-19 (SARS-CoV-2 coronavirus)  - Positive for influenza virus  - Positive for another respiratory virus  - Negative for all/any viruses tested  - Not Known |
| **Since you last checked in with us**, have you experienced any of the following symptoms: cold or flu symptoms, sore throat, persistent cough, loss of smell or taste, fever, fatigue, diarrhoea, abdominal pain or loss of appetite? | - Yes, I have had one or more of these symptoms since completing my last COVIDENCE UK questionnaire  - No, I have not had any of these symptoms since completing my last COVIDENCE UK questionnaire |
| When did your symptoms start? (DD/MM/YYYY) |  |
| Did you have a persistent cough (coughing a lot for more than an hour, or 3 or more coughing episodes in 24 hours)? | - No  - Persistent dry cough (i.e. producing little or no phlegm)  - Persistent productive cough |
| Did you experience unusual fatigue? | - No  - Mild fatigue  - Severe fatigue - I struggled to get out of bed |
| Did you have a loss of sense of smell or taste? | - Yes  - No |
| Did you skip any meals because you felt unwell? | - Yes  - No |

#### **Table S3.** Determinants of test-confirmed COVID-19: multivariable analysis excluding vaccinated participants (n=12,025)

| **Characteristic** | **Categories** | **Fully-adjusted odds ratio (95% CI)^1^** |
| --- | --- | --- |
| Age, years | 16-29.99 | 1.00 |
|  | 30-39.99 | 1.56 (0.8-3.04) |
|  | 40-49.99 | 1.59 (0.82-3.08) |
|  | 50-59.99 | 1.55 (0.81-2.96) |
|  | 60-69.99 | 1.30 (0.65-2.58) |
|  | ≥70 | 1.01 (0.46-2.19) |
| Sex | Female | 1.00 |
|  | Male | 0.98 (0.75-1.28) |
| Ethnicity | White | 1.00 |
|  | Mixed/Multiple/Other ethnic groups | 1.37 (0.76-2.47) |
|  | Asian/Asian British | 2.51 (1.37-4.58) |
|  | Black/African/Caribbean/Black British | 1.81 (0.67-4.93) |
| Highest educational level attained | Primary/Secondary | 1.00 |
|  | Higher/Further (A levels) | 0.97 (0.62-1.52) |
|  | College | 0.95 (0.65-1.40) |
|  | Post-graduate | 0.90 (0.60-1.36) |
| Index of multiple deprivation (IMD) rank, quartiles | Q1 | 1.08 (0.79-1.49) |
|  | Q2 | 0.89 (0.64-1.23) |
|  | Q3 | 0.85 (0.61-1.18) |
|  | Q4 | 1.00 |
| Housing | Owns own home | 1.00 |
|  | Mortgage | 1.29 (0.95-1.74) |
|  | Privately Renting | 1.03 (0.65-1.65) |
|  | Renting from council | 1.08 (0.57-2.06) |
|  | Others | 1.38 (0.78-2.42) |
| Number of people per bedroom | ≤0.50 | 1.00 |
|  | >0.50-0.99 | 1.25 (0.90-1.74) |
|  | 1-1.99 | 1.69 (1.22-2.33) |
|  | ≥2 | 2.39 (1.31-4.36) |
| Schoolchildren (aged 5-15 years) at home with participants | No | 1.00 |
|  | Yes | 1.33 (0.96-1.85) |
| Dog at home | No | 1.00 |
|  | Yes | 1.14 (0.88-1.47) |
| Shielding | No | 1.00 |
|  | Yes | 0.74 (0.40-1.37) |
| Any visits to/from other households in week prior to questionnaire completion | No | 1.00 |
|  | Yes | 1.42 (1.11-1.80) |
| Number of visits to shops and other indoor public places in week prior to questionnaire completion | Q1 | 1.00 |
|  | Q2 | 2.23 (1.36-3.68) |
|  | Q3 | 2.62 (1.56-4.40) |
|  | Q4 | 2.76 (1.63-4.67) |
| Travel to work/study in week prior to questionnaire completion | No | 1.00 |
|  | Yes | 1.38 (0.92-2.08) |
|  | Not currently working or studying | 1.26 (0.79-2.00) |
| Frontline worker | No | 1.00 |
|  | Health or social care worker | 1.45 (1.07-1.96) |
|  | Other frontline worker | 2.56 (1.71-3.83) |
| Number of portions of fruit, vegetables, and salad intake per day in week prior to questionnaire completion, quartiles | Q1 | 1.00 |
|  | Q2 | 1.07 (0.76-1.50) |
|  | Q3 | 1.11 (0.76-1.63) |
|  | Q4 | 0.91 (0.63-1.3) |
| Taking vitamin D supplement | No | 1.00 |
|  | Yes | 0.95 (0.74-1.21) |
| Hours of lower-impact physical activity in week prior to questionnaire completion^2^ | 0 | 1.00 |
|  | 1 | 0.99 (0.74-1.33) |
|  | ≥2 | 0.81 (0.60-1.10) |
| Estimated average hours of sleep per night in month prior to questionnaire completion^3^ | ≤6 | 1.17 (0.77-1.77) |
|  | 7 | 1.17 (0.87-1.57) |
|  | 8 | 1.00 |
|  | ≥9 | 1.29 (0.98-1.71) |
| Alcohol consumption in week prior to questionnaire completion, units | None | 1.00 |
|  | 1-7 | 1.18 (0.88-1.58) |
|  | 8-14 | 1.17 (0.83-1.65) |
|  | 15-21 | 1.49 (0.98-2.26) |
|  | 22-28 | 1.20 (0.66-2.18) |
|  | >28 | 0.85 (0.41-1.74) |
| Body mass index, kg/m^2^ | <25 | 1.00 |
|  | 25-30 | 1.37 (1.06-1.78) |
|  | >30 | 1.35 (1.00-1.83) |
| Asthma | No | 1.00 |
|  | Yes | 0.83 (0.55-1.26) |
| Atopic disease^4^ | No | 1.00 |
|  | Yes | 0.78 (0.59-1.03) |
| Systemic Immunosuppressants | No | 1.00 |
|  | Yes | 0.48 (0.19-1.22) |
| Inhaled corticosteroids | No | 1.00 |
|  | Yes | 0.76 (0.37-1.57) |
| Bronchodilators^5^ | No | 1.00 |
|  | Yes | 1.13 (0.60-2.15) |
| BCG-vaccinated | No | 1.00 |
|  | Yes | 1.30 (0.89-1.90) |
|  | Unsure | 1.11 (0.65-1.87) |
| Follow-up duration (day) |  | 1.00 (1.00-1.00) |
| Swab test frequency |  | 1.77 (1.64-1.90) |

Abbreviation: BCG, *M. bovis* Bacillus Calmette Guérin

1, adjusted for age, sex, duration of participation, test frequency, ethnicity, highest educational level attained, IMD rank, hours of sleep per night, housing, number of people per bedroom, presence of schoolchildren at home, dog at home, shielding, visits to/from other households, visits to shops and other indoor places, travel to work or study, frontline worker status, fruit, vegetable and salad intake, supplemental vitamin D intake, low-impact physical activity, alcohol intake, body mass index, history of asthma, history of atopic disease, use of systemic immunosuppressants, use of inhaled corticosteroids, use of inhaled bronchodilators and BCG vaccination status.

2, defined as exercise to improve flexibility or core strength such as yoga, tai chi or pilates, reported to the nearest hour

3, reported to nearest hour

4, defined by atopic eczema/dermatitis and/or hayfever/allergic rhinitis

5, defined as β-2-adrenoceptor agonists or anticholinergics

#### **Table S4.** Determinants of test-confirmed COVID-19: multivariable analysis excluding participants randomised to receive vitamin D supplementation in the CORONAVIT trial (n=11,414)

| **Characteristic** | **Categories** | **Fully-adjusted odds ratio (95% CI)^1^** |
| --- | --- | --- |
| Age, years | 16-29.99 | 1.00 |
|  | 30-39.99 | 1.02 (0.54-1.92) |
|  | 40-49.99 | 1.10 (0.59-2.04) |
|  | 50-59.99 | 0.90 (0.49-1.65) |
|  | 60-69.99 | 0.60 (0.31-1.16) |
|  | ≥70 | 0.58 (0.28-1.20) |
| Sex | Female | 1.00 |
|  | Male | 0.95 (0.72-1.24) |
| Ethnicity | White | 1.00 |
|  | Mixed/Multiple/Other ethnic groups | 1.25 (0.68-2.28) |
|  | South Asian | 2.24 (1.24-4.04) |
|  | Black/African/Caribbean/Black British | 1.80 (0.62-5.24) |
| Highest educational level attained | Primary/Secondary | 1.00 |
|  | Higher/Further (A levels) | 1.12 (0.71-1.76) |
|  | College | 0.86 (0.57-1.29) |
|  | Post-graduate | 0.82 (0.53-1.25) |
| Index of multiple deprivation (IMD) rank, quartiles | Q1 | 1.15 (0.83-1.59) |
|  | Q2 | 0.95 (0.68-1.32) |
|  | Q3 | 0.94 (0.67-1.30) |
|  | Q4 | 1.00 |
| Housing | Owns own home | 1.00 |
|  | Mortgage | 1.25 (0.92-1.69) |
|  | Privately Renting | 1.02 (0.64-1.64) |
|  | Renting from council | 1.04 (0.52-2.06) |
|  | Others | 1.13 (0.63-2.05) |
| Number of people per bedroom | ≤0.50 | 1.00 |
|  | >0.50-0.99 | 1.02 (0.73-1.41) |
|  | 1-1.99 | 1.47 (1.07-2.03) |
|  | ≥2 | 1.69 (0.88-3.25) |
| Schoolchildren (aged 5-15 years) at home with participants | No | 1.00 |
|  | Yes | 1.11 (0.80-1.55) |
| Dog at home | No | 1.00 |
|  | Yes | 1.29 (1.00-1.66) |
| Shielding | No | 1.00 |
|  | Yes | 0.93 (0.53-1.65) |
| Any visits to/from other households in week prior to questionnaire completion | No | 1.00 |
|  | Yes | 1.28 (1.00-1.63) |
| Number of visits to shops and other indoor public places in week prior to questionnaire completion | Q1 | 1.00 |
|  | Q2 | 1.79 (1.13-2.83) |
|  | Q3 | 2.20 (1.37-3.56) |
|  | Q4 | 2.36 (1.45-3.85) |
| Travel to work/study in week prior to questionnaire completion | No | 1.00 |
|  | Yes | 1.62 (1.04-2.54) |
|  | Not currently working or studying | 1.53 (0.93-2.55) |
| Frontline worker | No | 1.00 |
|  | Health or social care worker | 1.40 (1.01-1.93) |
|  | Other frontline worker | 1.34 (0.96-1.87) |
| Number of portions of fruit, vegetables, and salad intake per day in week prior to questionnaire completion, quartiles | Q1 | 1.00 |
|  | Q2 | 1.09 (0.77-1.55) |
|  | Q3 | 1.23 (0.83-1.82) |
|  | Q4 | 1.04 (0.72-1.50) |
| Taking vitamin D supplement | No | 1.00 |
|  | Yes | 0.91 (0.72-1.15) |
| Hours of lower-impact physical activity in week prior to questionnaire completion^2^ | 0 | 1.00 |
|  | 1 | 1.04 (0.77-1.39) |
|  | ≥2 | 0.86 (0.64-1.16) |
| Estimated average hours of sleep per night in month prior to questionnaire completion^3^ | ≤6 | 1.04 (0.67-1.62) |
|  | 7 | 1.32 (0.99-1.76) |
|  | 8 | 1.00 |
|  | ≥9 | 1.24 (0.93-1.65) |
| Alcohol consumption in week prior to questionnaire completion, units | None | 1.00 |
|  | 1-7 | 1.12 (0.83-1.50) |
|  | 8-14 | 1.08 (0.77-1.53) |
|  | 15-21 | 1.30 (0.85-1.98) |
|  | 22-28 | 1.44 (0.82-2.55) |
|  | >28 | 0.74 (0.33-1.67) |
| Body mass index, kg/m^2^ | <25 | 1.00 |
|  | 25-30 | 1.52 (1.17-1.97) |
|  | >30 | 1.39 (1.02-1.89) |
| Asthma | No | 1.00 |
|  | Yes | 0.92 (0.61-1.39) |
| Atopic disease^4^ | No | 1.00 |
|  | Yes | 0.77 (0.59-1.02) |
| Systemic Immunosuppressants | No | 1.00 |
|  | Yes | 0.31 (0.11-0.89) |
| Inhaled corticosteroids | No | 1.00 |
|  | Yes | 0.93 (0.47-1.83) |
| Bronchodilators^5^ | No | 1.00 |
|  | Yes | 0.99 (0.53-1.86) |
| BCG-vaccinated | No | 1.00 |
|  | Yes | 1.27 (0.87-1.86) |
|  | Unsure | 1.09 (0.64-1.85) |
| Follow-up duration (day) |  | 1.00 (1.00-1.01) |
| Swab test frequency |  | 1.58 (1.47-1.70) |

Abbreviation: BCG, *M. bovis* Bacillus Calmette Guérin

1, adjusted for age, sex, duration of participation, test frequency, ethnicity, highest educational level attained, IMD rank, hours of sleep per night, housing, number of people per bedroom, presence of schoolchildren at home, dog at home, shielding, visits to/from other households, visits to shops and other indoor places, travel to work or study, frontline worker status, fruit, vegetable and salad intake, supplemental vitamin D intake, low-impact physical activity, alcohol intake, body mass index, history of asthma, history of atopic disease, use of systemic immunosuppressants, use of inhaled corticosteroids, use of inhaled bronchodilators and BCG vaccination status.

2, defined as exercise to improve flexibility or core strength such as yoga, tai chi or pilates, reported to the nearest hour

3, reported to nearest hour

4, defined by atopic eczema/dermatitis and/or hayfever/allergic rhinitis

5, defined as β-2-adrenoceptor agonists or anticholinergics

#### **Table S5.** Association between asthma endotypes and test-confirmed COVID-19

| **Asthma** | **Atopy** | **Total number of participants (%)** | **Number of cases** | **Crude odds ratio (95% CI)** | **Minimally adjusted odds ratio (95% CI)^1^** | **Standard fully adjusted odds ratio (95% CI)^2^** | **Odds ratio (95% CI) additionally adjusted for inhaled corticosteroids** |
| --- | --- | --- | --- | --- | --- | --- | --- |
| No | No | 10167 (66.8) | 315 | 1.00 | 1.00 | 1.00 | 1.00 |
| No | Yes | 2564 (16.8) | 69 | 0.86 (0.66-1.13) | 0.72 (0.55-0.94) | 0.78 (0.59-1.03) | 0.78 (0.59-1.03) |
| Yes | No | 1163 (7.6) | 34 | 0.94 (0.66-1.35) | 0.80 (0.55-1.15) | 0.91 (0.62-1.33) | 0.92 (0.61-1.38) |
| Yes | Yes | 1333 (8.8) | 28 | 0.67 (0.45-0.99) | 0.49 (0.33-0.73) | 0.64 (0.42-0.96) | 0.65 (0.42-1.01) |

1, adjusted for age, sex, duration of participation and test frequency

2, adjusted for age, sex, duration of participation, test frequency and other covariables included in the final model for test-confirmed COVID-19 (i.e. ethnicity, highest educational level attained, IMD rank, hours of sleep, housing, number of people per bedroom, presence of schoolchildren at home, dog at home, shielding, visits to/from other households, visits to shops and other indoor places, travel to work or study, frontline worker status, fruit, vegetable and salad intake, supplemental vitamin D intake, low-impact physical activity, alcohol intake, body mass index, history of asthma, history of atopy, use of systemic immunosuppressants, and BCG vaccination status.)

3, adjusted for all factors listed in footnote 2 above, plus inhaled corticosteroids

#### **Table S6.** Determinants of symptom-defined probable COVID-19: minimally-adjusted and multivariable analyses

| **Variable label** | **Categories** | **Number (%) with incident symptom-defined probable COVID-19** | **Minimally-adjusted odds ratio (95% CI)^1^** | **Fully-adjusted odds ratio (95% CI)^2^** | **P for trend** |
| --- | --- | --- | --- | --- | --- |
| Age, years | 16-29.99 | 42 (7.3) | 1.00 |  | <0.001 |
|  | 30-39.99 | 61 (6.2) | 0.83 (0.55-1.25) | 0.72 (0.45-1.16) |  |
|  | 40-49.99 | 112 (6.3) | 0.82 (0.57-1.19) | 0.73 (0.46-1.15) |  |
|  | 50-59.99 | 132 (3.9) | 0.50 (0.35-0.71) | 0.59 (0.37-0.92) |  |
|  | 60-69.99 | 120 (2.4) | 0.30 (0.21-0.43) | 0.50 (0.30-0.80) |  |
|  | ≥70 | 44 (1.3) | 0.17 (0.11-0.26) | 0.28 (0.16-0.49) |  |
| Sex | Female | 385 (3.6) | 1.00 |  | -- |
|  | Male | 126 (2.7) | 0.94 (0.76-1.15) | 0.90 (0.73-1.13) |  |
| Ethnicity | White | 470 (3.3) | 1.00 |  | -- |
|  | Mixed/Multiple/Other ethnic groups | 18 (4.5) | 1.04 (0.64-1.69) | 1.07 (0.65-1.76) |  |
|  | Asian/Asian British | 21 (7.5) | 1.71 (1.08-2.72) | 1.85 (1.13-3.03) |  |
|  | Black/African/Caribbean/Black British | 2 (2.1) | 0.50 (0.12-2.03) | 0.53 (0.13-2.19) |  |
| Highest educational level attained | Primary/Secondary | 59 (3.6) | 1.00 |  | 0.84 |
|  | Higher/Further (A levels) | 75 (3.4) | 0.80 (0.56-1.13) | 0.84 (0.59-1.20) |  |
|  | College | 220 (3.3) | 0.77 (0.57-1.04) | 0.88 (0.65-1.20) |  |
|  | Post-graduate study | 157 (3.4) | 0.74 (0.54-1.01) | 0.91 (0.66-1.27) |  |
| Index of multiple deprivation (IMD) rank, quartiles | Q1 (most deprived) | 159 (4.4) | 1.33 (1.02-1.72) | 1.17 (0.90-1.54) | 0.15 |
|  | Q2 | 140 (3.7) | 1.30 (1.00-1.69) | 1.19 (0.91-1.57) |  |
|  | Q3 | 108 (2.8) | 1.05 (0.79-1.38) | 1.02 (0.77-1.35) |  |
|  | Q4 (least deprived) | 100 (2.6) | 1.00 | 1.00 |  |
| Claiming universal credit | No | 481 (3.3) | 1.00 |  | -- |
|  | Yes | 28 (6.9) | 1.51 (1.01-2.26) | 1.21 (0.80-1.85) |  |
| Housing | Owns own home | 199 (2.1) | 1.00 |  | -- |
|  | Mortgage | 195 (5.2) | 1.50 (1.17-1.91) | 1.35 (1.05-1.74) |  |
|  | Privately Renting | 53 (5.0) | 1.40 (0.98-1.99) | 1.15 (0.79-1.67) |  |
|  | Renting from council | 26 (5.5) | 1.94 (1.26-3.00) | 1.14 (0.71-1.86) |  |
|  | Others | 38 (6.2) | 1.60 (1.04-2.46) | 1.22 (0.77-1.93) |  |
| Number of people per bedroom | ≤0.5 | 119 (2.0) | 1.00 |  | 0.002 |
|  | >0.5-0.99 | 116 (2.8) | 1.14 (0.88-1.49) | 1.07 (0.81-1.40) |  |
|  | 1.00-1.99 | 249 (5.2) | 1.68 (1.32-2.15) | 1.45 (1.12-1.89) |  |
|  | ≥2 | 24 (7.2) | 2.25 (1.39-3.62) | 1.75 (1.06-2.91) |  |
| Pre-school children (age 0-4 years) at home with participant | No | 470 (3.2) | 1.00 |  | -- |
|  | Yes | 41 (8.1) | 1.51 (1.05-2.19) | 1.44 (0.98-2.13) |  |
| Schoolchildren (age 5-15 years) at home with participant | No | 398 (3.0) | 1.00 |  | -- |
|  | Yes | 113 (6.6) | 1.40 (1.09-1.80) | 1.19 (0.91-1.56) |  |
| Dog at home | No | 353 (3.0) | 1.00 |  | -- |
|  | Yes | 158 (4.4) | 1.30 (1.07-1.58) | 1.22 (1.00-1.50) |  |
| Shielding | No | 462 (3.3) | 1.00 |  | -- |
|  | Yes | 49 (4.2) | 1.32 (0.97-1.79) | 0.89 (0.62-1.29) |  |
| Frontline worker | No | 368 (3.0) | 1.00 |  | -- |
|  | Health or social care worker | 50 (4.3) | 0.98 (0.72-1.33) | 0.98 (0.71-1.36) |  |
|  | Other frontline worker | 92 (5.9) | 1.61 (1.26-2.05) | 1.51 (1.17-1.94) |  |
| Number of portions of fruit, vegetables, and salad intake per day in week prior to questionnaire completion, quartiles | Q1 | 97 (4.4) | 1.00 |  | 0.25 |
|  | Q2 | 178 (3.5) | 0.84 (0.65-1.08) | 0.91 (0.70-1.19) |  |
|  | Q3 | 80 (2.8) | 0.67 (0.49-0.91) | 0.74 (0.54-1.02) |  |
|  | Q4 | 154 (3.0) | 0.79 (0.61-1.03) | 0.87 (0.65-1.15) |  |
| Portions of dairy products per day in week prior to questionnaire completion, quartiles | 0-1/d | 135 (3.3) | 1.00 |  | 0.10 |
|  | 2/d | 141 (3.2) | 1.06 (0.83-1.35) | 1.11 (0.86-1.42) |  |
|  | 3-5/d | 128 (3.5) | 1.20 (0.94-1.54) | 1.23 (0.95-1.58) |  |
|  | 6+/d | 106 (3.4) | 1.28 (0.98-1.67) | 1.22 (0.92-1.62) |  |
| Vitamin C supplement | No | 446 (3.3) | 1.00 |  | -- |
|  | Yes | 65 (4.2) | 1.27 (0.97-1.66) | 1.25 (0.95-1.65) |  |
| Cod liver oil supplement | No | 464 (3.3) | 1.00 |  | -- |
|  | Yes | 47 (3.8) | 1.47 (1.08-2.01) | 1.52 (1.10-2.09) |  |
| Estimated average hours of sleep per night in month prior to questionnaire completion^3^ | ≤6 | 60 (4.6) | 1.61 (1.19-2.18) | 1.25 (0.91-1.73) | 0.10^4^ |
|  | 7 | 120 (3.3) | 1.16 (0.92-1.47) | 1.09 (0.86-1.39) |  |
|  | 8 | 179 (2.9) | 1.00 | 1.00 |  |
|  | ≥9 | 152 (3.8) | 1.17 (0.93-1.46) | 1.15 (0.91-1.45) |  |
| Hours of vigorous physical exercise in week prior to questionnaire completion^5^ | 0 | 216 (3.7) | 1.00 |  | 0.94 |
|  | 1-3 | 188 (3.3) | 0.91 (0.75-1.12) | 1.05 (0.85-1.31) |  |
|  | ≥4 | 105 (2.8) | 0.79 (0.62-1.00) | 1.00 (0.77-1.29) |  |
| Tobacco smoking status | Never-smoker | 274 (3.2) | 1.00 |  | -- |
|  | Ex-smoker | 200 (3.4) | 1.21 (1.00-1.47) | 1.03 (0.84-1.27) |  |
|  | Current smoker | 37 (4.4) | 1.19 (0.83-1.69) | 0.84 (0.56-1.26) |  |
| Vaping status | Never-vaper | 454 (3.2) | 1.00 |  | -- |
|  | Ex-vaper | 23 (4.6) | 1.15 (0.74-1.77) | 0.96 (0.59-1.56) |  |
|  | Current vaper | 33 (6.9) | 1.68 (1.16-2.44) | 1.37 (0.91-2.07) |  |
| Self-rated general physical health | Excellent | 78 (2.5) | 1.00 |  | 0.002 |
|  | Very good | 172 (2.9) | 1.17 (0.89-1.53) | 1.12 (0.85-1.49) |  |
|  | Good | 149 (3.7) | 1.50 (1.13-1.99) | 1.29 (0.95-1.75) |  |
|  | Fair | 84 (5.1) | 2.00 (1.46-2.75) | 1.67 (1.16-2.40) |  |
|  | Poor | 28 (6.2) | 2.26 (1.44-3.54) | 1.77 (1.06-2.97) |  |
| Feeling anxious or depressed today | No | 314 (2.8) | 1.00 |  | -- |
|  | Yes | 197 (4.8) | 1.42 (1.18-1.72) | 1.12 (0.90-1.38) |  |
| Body mass index, kg/m^2^ | <25 | 226 (3.0) | 1.00 |  | 0.89 |
|  | 25-30 | 165 (3.4) | 1.23 (1.00-1.51) | 1.13 (0.91-1.40) |  |
|  | >30 | 120 (4.1) | 1.29 (1.02-1.62) | 0.99 (0.77-1.27) |  |
| Proton pump inhibitors | No | 438 (3.3) | 1.00 |  | -- |
|  | Yes | 73 (3.5) | 1.31 (1.01-1.69) | 0.97 (0.73-1.30) |  |
| Selective serotonin reuptake inhibitors | No | 442 (3.1) | 1.00 |  | -- |
|  | Yes | 69 (6.4) | 1.70 (1.30-2.22) | 1.41 (1.06-1.89) |  |
| Paracetamol | No | 480 (3.3) | 1.00 |  | -- |
|  | Yes | 31 (4.8) | 1.79 (1.23-2.61) | 1.28 (0.84-1.94) |  |
| Systemic immunosuppressants | No | 480 (3.3) | 1.00 |  | -- |
|  | Yes | 31 (4.5) | 1.38 (0.95-2.01) | 1.08 (0.70-1.67) |  |
| Inhaled bronchodilators^6^ | No | 448 (3.2) | 1.00 |  | -- |
|  | Yes | 63 (4.5) | 1.26 (0.96-1.66) | 1.12 (0.83-1.50) |  |
| Follow-up duration (days) |  |  |  | 1.00 (1.00-1.01) | -- |

1, adjusted for age, sex and duration of participation

2, adjusted for age, sex, duration of participation, ethnicity, highest educational level attained, IMD rank, claiming universal credit, housing, number of people per bedroom, presence of pre-school children at home, presence of schoolchildren at home, dog at home, shielding, frontline worker status, fruit, vegetable and salad intake, dairy product intake, supplemental vitamin C intake, cod liver oil intake, hours of sleep per night, vigorous physical exercise, smoking, vaping, general physical health, feeling anxious or depressed, body mass index, and use of proton pump inhibitors, SSRIs, paracetamol, systemic immunosuppressants and bronchodilators.

3, reported to the nearest hour

4, only applies to sleep categories below 8 hours

5, defined as exercise of sufficient intensity to make the participant breathless or to raise their heart rate significantly, such as heavy physical work, strenuous gardening (e.g. vigorous digging, landscaping) swimming, jogging, aerobics, football, tennis, cycling, gym workout, reported to the nearest hour

6, inhaled β-2-adrenoceptor agonists or anticholinergics

### **Supplementary Figures**

#### **Figure S1.** Participant Flow

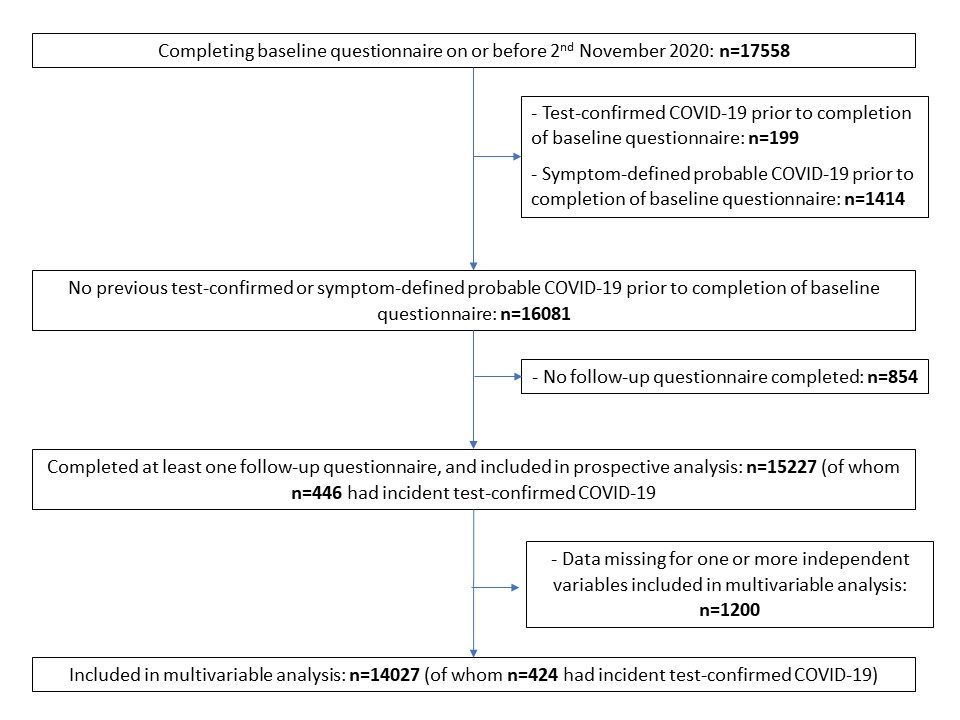

#### **Figure S2.** Heat-map illustrating geographical distribution of COVIDENCE UK participants (A) and incidence of COVID-19 (B) in the UK. Data for (B) from Public Health England, 2020.

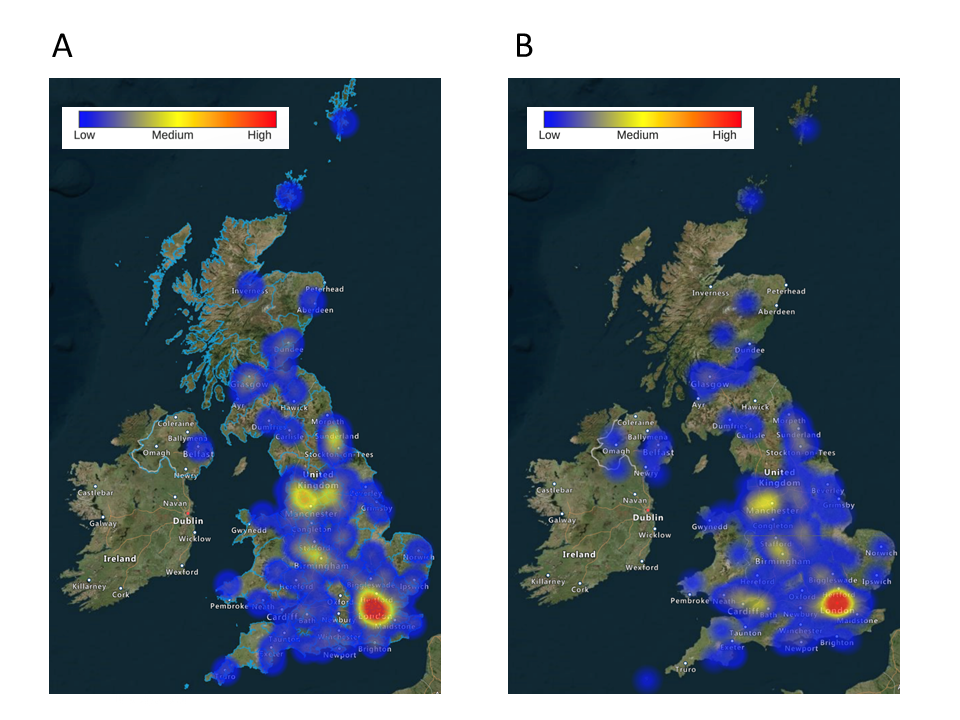

#### **Figure S3.** Heat-map illustrating degree of collinearity between independent variables associating with test-confirmed COVID-19 risk in minimally adjusted model.

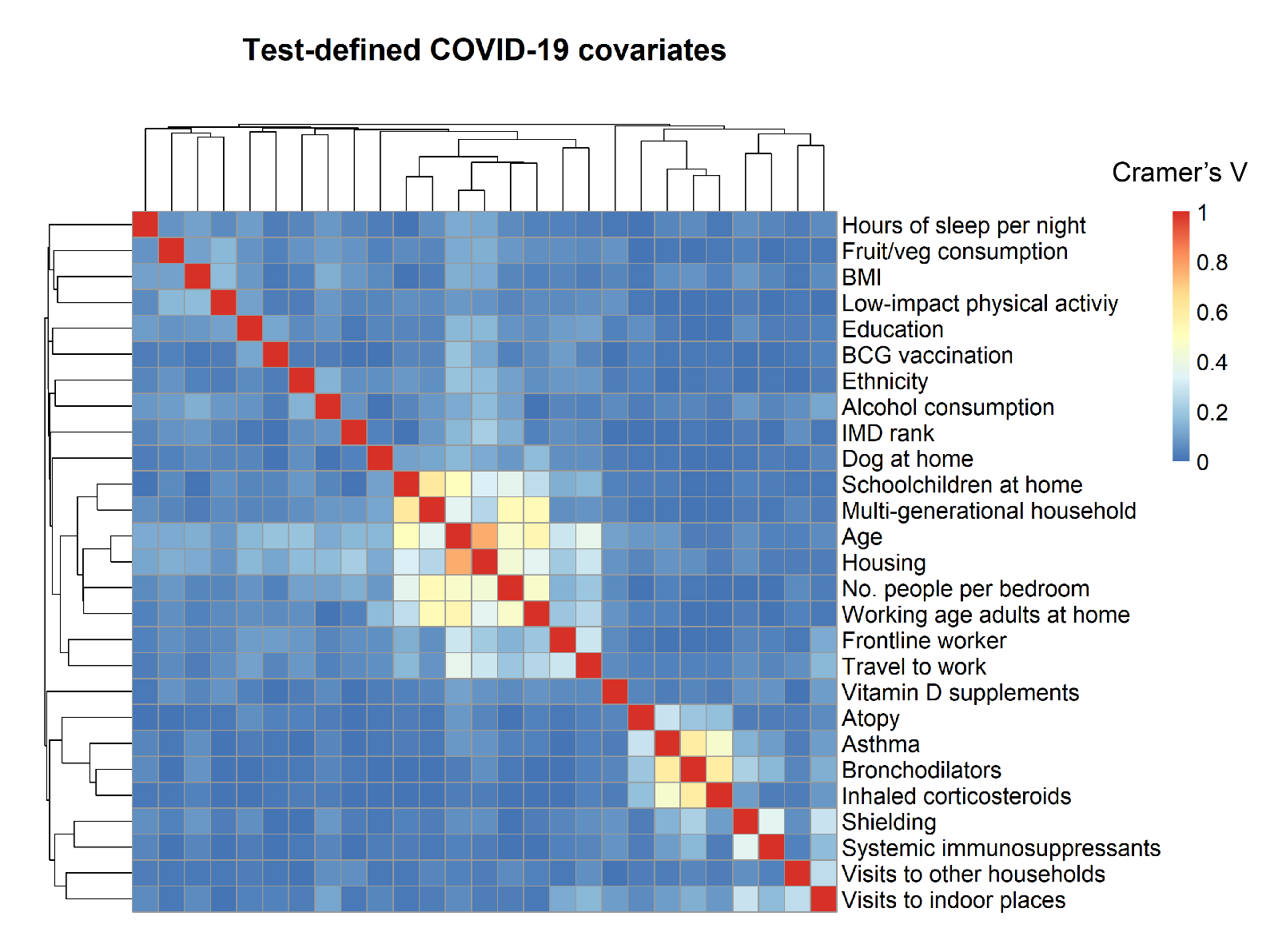
